# Multidomain residential external exposome and epigenetic ageing in the UK general population

**DOI:** 10.64898/2026.09.15.26363118

**Authors:** Gergő Baranyi, Kees de Hoogh, Fabián Coloma, Roel Vermeulen, Meena Kumari, Cathryn Tonne

## Abstract

Residential environmental exposures might be associated with biological ageing, but evidence has largely focused on a limited set of single exposures. We investigated associations between the multidomain residential external exposome and epigenetic ageing in 3307 adults from the nationally representative UK Understanding Society study. We linked 30 exposures across six domains spanning social, built, physico-chemical, retail, health and weather environments, to residential postcodes and examined associations with first-, second- and third-generation epigenetic clocks. Across four complementary approaches capturing individual, joint, domain-level and clustered exposures, adverse social, built and physico-chemical environments showed the most consistent associations with biological ageing. Area deprivation, neighbourhood crime, lower social cohesion and greenness, air pollution and higher temperatures were associated with faster pace of ageing measured by third-generation clocks (particularly DunedinPACE) and, to a lesser extent, with PhenoAge. Greater distance to leisure services was associated with older biological age on Horvath’s clock and PhenoAge. Clustering identified urban and rural exposure profiles, with faster ageing in the urban profile. Residential environmental exposures are associated with epigenetic ageing, with later-generation clocks appearing more sensitive to the biological imprint of the external residential exposome.

**Graphical abstract:** 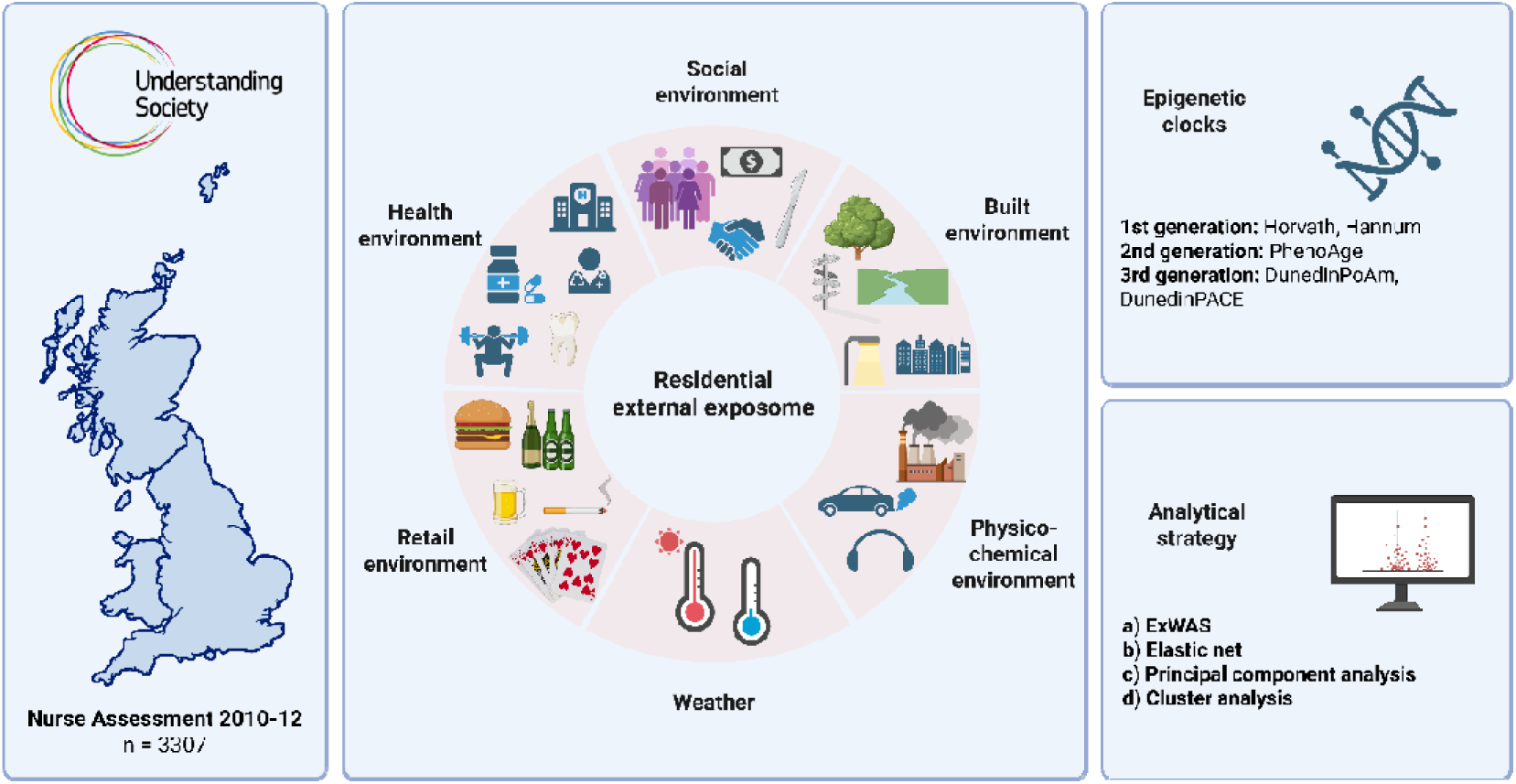

## INTRODUCTION

Environmental conditions, including air pollution, noise and temperature, are major determinants of population health, ^1^ yet the processes through which they become biologically embedded are not fully understood. Epigenetic alterations, including DNA methylation, represent one potential mechanistic link between environmental exposures and health outcomes^2^. DNA methylation involves the addition of a methyl group to cytosine– phosphate–guanine (CpG) dinucleotides, which regulates gene expression without altering the underlying DNA sequence^3^. DNA methylation patterns change with age, and methylation levels at specific CpG sites can accurately predict chronological age^4^.

Based on age-related methylation patterns, epigenetic clocks have been developed using machine learning approaches to estimate biological age^5^. Whereas first-generation clocks, including Horvath’s^6^ and Hannum’s^7^, were trained to predict chronological age, second-generation clocks, such as PhenoAge^8^, were developed to better predict survival by incorporating clinical biomarkers^5^. The deviation between biological and chronological age (i.e., age acceleration) has been linked to mortality^9^ and various diseases^4,10^. In contrast, third-generation clocks quantify the pace rather than the level of ageing and were trained on longitudinal changes in biomarkers measured repeatedly within the same cohort over time^11^.

Existing evidence linking environmental conditions to epigenetic ageing has focused largely on single exposures, particularly air pollution^12^ and neighbourhood deprivation^13^. While social, built, physico-chemical and service environments simultaneously affect health, studies rarely consider multiple environmental factors at the same time^14,15^. The exposome framework provides a useful approach^16^ and novel methodologies^17,18^ for moving beyond traditional single-exposure studies by characterising the complex, multidimensional environmental conditions in which people live. This framework can help identify which environmental dimensions and mixtures are most consistently associated with epigenetic ageing, and whether later-generation clocks are more sensitive to environmental exposures than their predecessors. Finally, well-characterised population-based cohort data are crucial to robust adjustment for confounders, producing generalisable findings, and capturing often overlooked population groups, including rural populations.

We investigated how the residential external exposome spanning multiple domains relates to epigenetic ageing in 3307 UK adults from the nationally representative Understanding Society study. Thirty exposures across six domains (social, built, physico-chemical, retail, health and weather) were linked to residential postcodes and their associations with first-, second- and third-generation epigenetic clocks were examined. By combining single-exposure models, penalised regression, domain-level dimension reduction and clustering approaches, we aimed to identify the environmental features most consistently associated with biological ageing.

## METHODS

Understanding Society, the UK Household Longitudinal Study (UKHLS) is a large, nationally representative longitudinal panel survey of UK households established in 2009. The study annually follows approximately 40000 households, including surviving members of the British Household Panel Survey who were incorporated into the sample in 2010^19,20^. In waves 2 and 3 (2010–2012), adult participants who consented to a health assessment were visited by a nurse approximately five months after the main interview. The assessment included detailed physical health measurements (e.g., anthropometric measures, blood pressure) and the collection of a blood sample. Participants were eligible if they were aged ≥16 years, resided in Great Britain (England, Wales, or Scotland), completed the main interview in English, and were not pregnant. ^19^ Among eligible participants, 57.6% took part in the nurse assessment, and 63.4% of those participants consented to and provided a blood sample (∼13000) ^21^. Ethical approval was obtained from the National Research Ethics Service (10/H0604/2).

### Epigenetic clocks

DNA methylation profiles were obtained from whole blood for 3654 participants of White ethnicity, including 1425 samples from wave 2 and 2229 from wave 3^22^. Profiling was conducted in two batches (2017: n=1174; 2020: n=2480), using the Illumina Methylation EPIC BeadChip, which measures over 850000 DNA methylation sites across the genome. Quality control procedures included the removal of outliers, filtering of poor-quality probes (<85% bisulfite conversion), and exclusion of samples showing large discrepancies between raw and normalised methylation values^23^. Five established epigenetic clocks were used: the first-generation Horvath’s multi-tissue^6^, and Hannum’s clocks^7^, the second-generation PhenoAge^8^, and the third-generation DunedinPoAm^11^, and DunedinPACE^24^. Whereas Horvath’s and Hannum’s clocks were trained to predict chronological age, PhenoAge was designed to better capture phenotypic ageing using nine clinical biomarkers^8^. DunedinPoAm and DunedinPACE were derived from longitudinal biomarker change and quantify the pace, rather than the level, of biological ageing, with the updated DunedinPACE being trained on 20 years of follow-up data capturing the ongoing rate of decline in system integrity. First- and second-generation clocks are expressed in years, whereas third-generation clocks as years of biological ageing per calendar year.

### Environmental domains

Participants’ residential addresses at the time of interview were used to link 30 exposures across six domains to the sample^25^. Environmental features available as high-resolution raster data from the EXPANSE repository^26^ were linked to residential postcode centroids (approximately 16 households), deriving area-weighted average exposure within 300m, 500m, and 1000m buffers. Other open data (i.e., Access to Healthy Assets and Hazards [AHAH] dataset^27^, English/Scottish/Welsh Index of Multiple Deprivation) were only available at LSOA level (i.e., Lower Layer Super Output Area, approximately 400–1200 households) and we used look-up tables to assign these to postcodes.

*Social environment* included area-level income deprivation (deciles from English/Scottish/Welsh Index of Multiple Deprivation), crime (deciles from English/Scottish/Welsh Index of Multiple Deprivation), social cohesion (shortened version of Buckner’s Neighbourhood Cohesion^28^ scale aggregated at LSOA level for all UKHLS respondents between 2008-2012) and population density (from the Global Human Settlement population grid) ^26^. *Built environment* included greenness (measured using the Normalized Difference Vegetation Index) ^26^, distance to green space (Euclidean distance to nearest accessible green land) ^26^, distance to blue space (Euclidean distance to nearest sea or inland freshwater body) ^26^, walkability (composite index based on factors related to walking) ^26^, light at night (night-time light intensity or radiance) ^26^, grey space (artificial or built-up land cover) ^26^, and geographic accessibility (deciles from English/Scottish/Welsh Index of Multiple Deprivation). Information on retail and health services were from the AHAH dataset^27^. *Retail environment* captured road network distance in kilometres to the nearest fast-food outlets, pubs, off-licences, tobacconists, and gambling outlets; business addresses were based on data from the Local Data Company^27^. The *health environment* captured road network distance in kilometres to the nearest general practitioners (GPs), hospitals, dentists, pharmacies, and leisure services based on data from National Health Service and the Local Data Company^27^. The *physico-chemical environment* domain included average day-evening-night road traffic noise (L_den_) for agglomerations (>100,000 people) and major roads (>3m vehicle annually) using government-reported estimates produced in accordance with the Environmental Noise Directive^29^, as well as rolling 12-months concentration of PM_2.5_, PM_10_, NO_2_, and O_3_ prior to the interview averaged from monthly EXPANSE estimates^26^. Finally, *weather* variables were computed as the mean and standard deviation (SD) of summer (June-August) and winter (December-February) temperatures during the 12 months preceding the interview, derived from EXPANSE^26^. Detailed description and sources of all variables can be found in Table S1.

### Covariates

Factors likely confounding the relationship between environmental exposures and epigenetic clocks were extracted from the main survey and are presented in a directed acyclic graph (Figure S1). They included age (continuous), sex (male, female), data collection wave (wave2, wave3), household income quartile (Q1–Lowest, Q2, Q3, Q4–Highest), country of birth (UK, non-UK), housing tenure (owned, private rent, social rent, other), partnership status (living alone, living with partner), economic activity (employed, long-term sick/disabled, maternity/caring, retired, unemployed, student/other) and government region. To account for blood cell composition and processing batch effects, we included white blood cell counts (CD8T, CD4T, NK, B cell, monocyte, and granulocyte) derived using Houseman’s algorithm^30^ and barcode information. To reduce multicollinearity, these technical variables were summarised into a single component using Factor Analysis of Mixed Data.

In a sensitivity analysis, we also considered smoking status (current, former, never), moderate physical activity (weekly, monthly, less frequent, not active) and body mass index (<18.5, 18.5 to <25.0, 25.0 to <30.0, 30.0 to <40.0, ≥40); however, they likely mediate the association between some environmental exposures and epigenetic clocks, and should therefore be interpreted cautiously.

### Statistical analysis

Environmental exposures were analysed as continuous variables and rescaled prior to modelling by dividing each by its interquartile range (IQR; Table S2). Survey weights were applied to account for unequal selection probabilities and non-response. Multicollinearity among exposures and confounders was assessed using the variance inflation factor (VIF).

To capture relevant features of the multidimensional residential external exposome, we used four complementary analytical approaches. First, we fitted weighted linear regression models separately for each exposure (exposure-wide association study; ExWAS) to estimate individual exposure–clock associations. False discovery rate correction^31^ was applied across exposures to account for multiple testing.

Second, Gaussian elastic net regression was used to examine the joint associations between correlated environmental exposures and biological ageing. Elastic net is a penalised regression approach that combines L1 (least absolute shrinkage and selection operator [LASSO]) and L2 (ridge regression) penalties. ^32^ This approach enables variable selection while accounting for multicollinearity among correlated exposures by shrinking less informative exposures towards zero and retaining predictors with stronger contributions. The elastic net mixing parameter (α), which determines the balance between L1 and L2 penalisation, and regularisation parameter (λ), which controls the degree of coefficient shrinkage, were selected using repeated 5-fold cross-validation to ensure both model stability and predictive performance. The optimal α was chosen based on the lowest cross-validated prediction error, while λ was selected as λ-min, corresponding to the value that minimised the cross-validated prediction error. Scaled exposure variables were entered simultaneously into the models alongside confounders (categorical variables were entered as dummy variables) and the technical adjustment variable: environmental exposures were penalised, whereas covariates were retained in all models by assigning them a penalty factor of zero.

Third, we assessed whether exposures within each environmental domain could be summarised by a common latent dimension using exploratory factor analysis. Domain suitability was evaluated using the Kaiser–Meyer–Olkin measure of sampling adequacy (reflecting the proportion of shared variance) and Bartlett’s test of sphericity (testing whether the correlation matrix deviated from an identity matrix). We then inspected factor loadings and explained variance to determine whether a dominant unidimensional structure was supported. For domains meeting these criteria, we derived a domain-level summary score using the first principal component, which captured the largest share of common variation across exposures and provided a parsimonious composite measure for use in weighted linear regression models of biological ageing.

Fourth, K-means clustering was used to identify groups of individuals with similar environmental exposure profiles after scaling input variables. The algorithm partitions observations into K mutually exclusive clusters by minimising within-cluster variation and maximising between-cluster separation, such that observations within clusters are as similar as possible and observations between clusters as distinct as possible. ^33^ We used three methods to determine the optimal number of clusters: i) the elbow method based on within-cluster sum of squares, ii) the silhouette method assessing the similarity of observations to their assigned cluster relative to other clusters, and (iii) the gap statistic comparing observed within-cluster variation to that expected under a reference null distribution. After identifying the optimal number of clusters, K-means clustering was conducted with 25 random initializations to improve solution stability; cluster membership was subsequently included as the exposure in weighted linear regression models, estimating its associations with biological ageing.

Four sensitivity analyses were conducted and presented alongside the main findings. First, models were additionally adjusted for smoking status, moderate physical activity, and body mass index. Second, missing data (9%) were imputed via multiple imputation by chained equations (10 datasets) with estimates pooled using Rubin’s rules. Third, in ExWAS we used 300m buffers in the main analysis, with 500m and 1000m buffers examined in sensitivity analyses. Fourth, we fitted mutually adjusted models including all domain-level summary scores simultaneously.

All analyses were conducted using the *survey, glmnet, factoextra, psych, mice* and *stats* packages in R 4.3.0. ^34^

## RESULTS

Out of 3654 participants with epigenetic clocks, 3307 (91%) were included in our final sample after dropping participants with missing exposure, confounder and survey weight data (Figure S2). Approximately 54% of the sample were female and 60% employed. The average weighted chronological age was 49 years, while biological ages were 55, 48 and 42 years based on the Horvath, Hannum and PhenoAge clocks; for DunedinPoAm and DunedinPACE the weighted mean was 1.0 (Table 1). Correlation between the 30 exposures is presented in Figure S3, and estimates are expressed as interquartile range (IQR) change.

**Table 1.** Descriptive characteristics of the analytical sample.

| Variables | Analytical sample<br>(n=3307) |
| --- | --- |
| Sex, n (%) |  |
| Male | 1457 (45.6%) |
| Female | 1850 (54.4%) |
| Age, mean ± SD | 48.9 ± 16.8 |
| Data collection wave, n (%) |  |
| Wave 2 | 2067 (72.5%) |
| Wave 3 | 1240 (27.5%) |
| Household income quartile, n (%) |  |
| Q1 - Lowest | 809 (25.8%) |
| Q2 | 818 (24.7%) |
| Q3 | 844 (25.7%) |
| Q4 - Highest | 836 (23.8%) |
| Country of birth, n (%) |  |
| UK | 3177 (94.6%) |
| Not UK | 130 (5.4%) |
| Housing tenure, n (%) |  |
| Owned | 2650 (72.3%) |
| Private rent | 249 (11.5%) |
| Social rent, Other | 408 (16.2%) |
| Partnership status, n (%) |  |
| Living with partner | 2413 (68.5%) |
| Living alone | 894 (31.5%) |
| Economic activity, n (%) |  |
| Employed | 1897 (60.0%) |
| Long-term sick/disabled | 104 (3.7%) |
| Maternity/Caring | 161 (5.3%) |
| Retired | 980 (23.1%) |
| Unemployed | 110 (5.3%) |
| Student/other | 55 (2.6%) |
| Smoking status, n (%) |  |
| Current smoker | 625 (23.1%) |
| Former smoker | 1340 (37.4%) |
| Never smoker | 1342 (39.5%) |
| Moderate physical activity, n (%) |  |
| Weekly | 1187 (35.5%) |
| Monthly | 425 (13.8%) |
| Less frequent | 568 (17.5%) |
| Not active | 1127 (33.2%) |
| Body Mass Index, n (%) |  |
| <18.5 | 27 (1.3%) |
| 18.5 to <25.0 | 952 (30.7%) |
| 25.0 to <30.0 | 1290 (37.6%) |
| 30.0 to <40.0 | 947 (27.4%) |
| $\geq 40.0$ | 91 (2.9%) |
| Horvath clock, mean ± SD | 55.0 ± 11.5 |
| Hannum clock, mean ± SD | 48.0 ± 12.1 |
| PhenoAge, mean ± SD | 42.3 ± 13.7 |
| DunedinPoAm, mean ± SD | 1.0 ± 0.1 |
| DunedinPACE, mean ± SD | 1.0 ± 0.1 |

Unweighted frequencies and weighted percentages are presented for categorical variables; weighted means and standard deviations (SD) are presented for continuous variables.

### Single exposure models with ExWAS

After adjusting for confounders and technical variables, and considering multiple testing, 1-IQR increase in distance to gambling outlets (b=0.035, 95%CI: 0.022, 0.048) and leisure services (b=0.060, 95%CI: 0.030, 0.091) was associated with Horvath’s clock and with PhenoAge (gambling outlets: b=0.036 [95%CI: 0.021, 0.050]; leisure services: b=0.066 [95%CI: 0.035, 0.097]). Area deprivation (b=0.007, 95%CI: 0.003, 0.012), crime (b=0.007 95%CI: 0.003, 0.011), less greenness (b=−0.006, 95%CI: −0.009, −0.002) and higher summer temperature (b=0.009, 95%CI: 0.003, 0.014) was associated with faster pace of ageing using DunedinPoAm. Area deprivation (b=0.013, 95%CI: 0.005, 0.020), crime (b=0.011, 95%CI: 0.004, 0.019), lower social cohesion (b=−0.009, 95%CI: −0.014, −0.003), higher population density (b=0.010, 95%CI: 0.004, 0.017), lower greenness (b=−0.012, 95%CI: −0.018, −0.006), higher walkability (b=0.009, 95%CI: 0.001, 0.016), more grey space (b=0.013, 95%CI: 0.006, 0.021), higher PM_2.5_ (b=0.018, 95%CI: 0.010, 0.027), PM_10_ (b=0.018, 95%CI: 0.010, 0.027), NO_2_ (b=0.013, 95%CI: 0.006, 0.021), and lower O_3_ (b=−0.014, 95%CI: −0.023, −0.005), and higher winter temperature (b=0.010, 95%CI: 0.002, 0.018) were associated with faster pace of ageing using DunedinPACE (Figure 1). In the sensitivity analysis after including smoking, physical activity and BMI, findings for Horvath’s clock and PhenoAge remained significant. For DunedinPoAm, summer temperature, for DunedinPACE social cohesion, greenness and air pollution (PM_2.5_, PM_10_, NO_2_, O_3_) remained significant (Tables S3-S7). Multiple imputation produced almost identical findings (Figure S4). Using larger buffer sizes (500m, 1000m) led to marginal differences in the estimates; with the exception of PhenoAge where greenness and NO_2_ became significant using the 1000m buffer (Figure S5).

**Figure 1.**
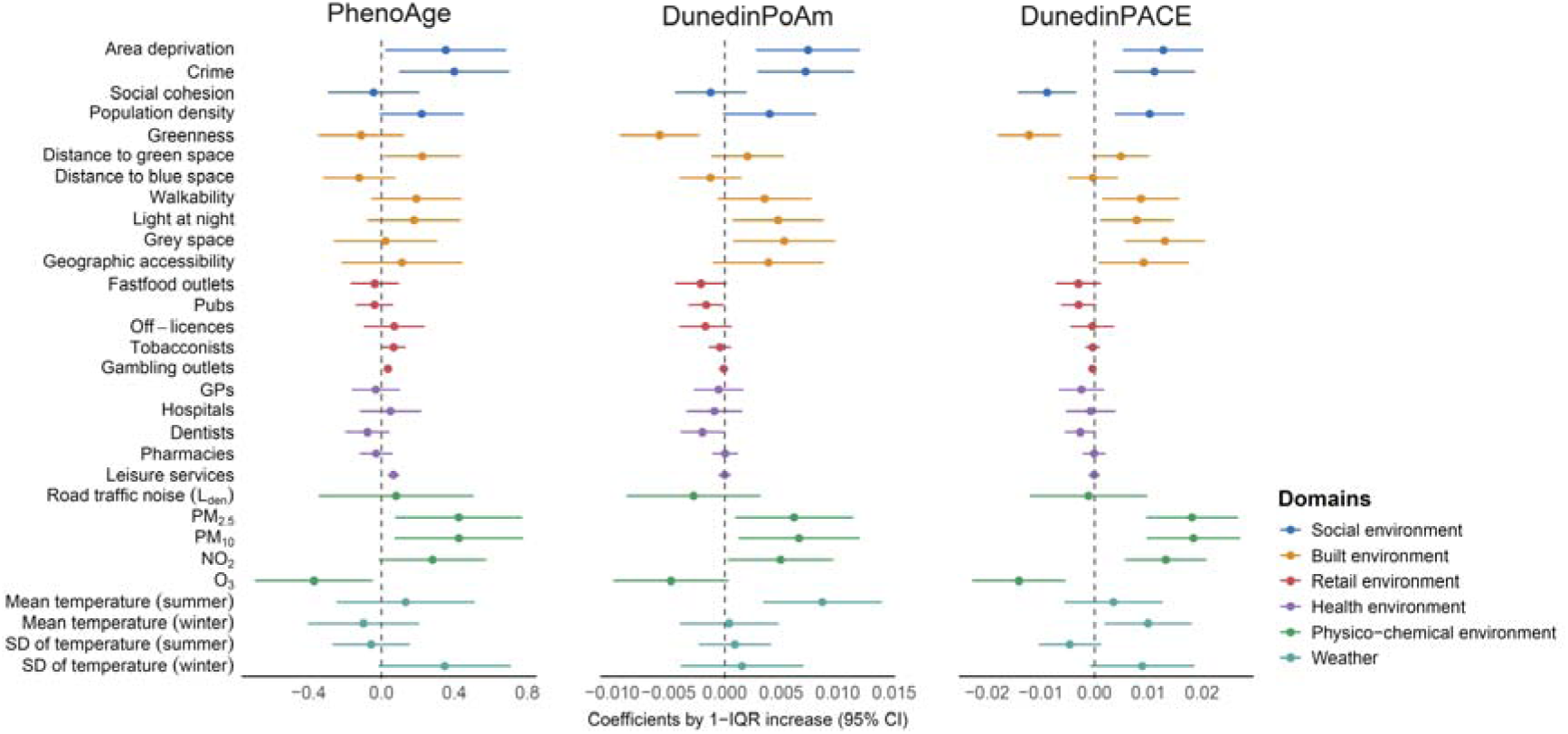
Associations of individual environmental exposures with epigenetic clocks. Weighted linear regression models were fitted separately for each exposure–clock combination and estimates are expressed per interquartile range (IQR) increase in exposure. Models were adjusted for age, sex, data collection wave, household income quartile, country of birth, housing tenure, partnership status, economic activity, government region, and technical variables summarised as a single component. Sample size was n = 3307.

### Variable selection with elastic net

Elastic net selected leisure services for Horvath’s and crime for Hannum’s clock, although with near-zero coefficients (b<0.001). For PhenoAge, crime (b=0.179) and leisure services (b=0.001), for DunedinPoAm area deprivation (b=0.003), crime (0.003), and greenness (b=−0.002) were selected. For DunedinPACE, area deprivation (b=0.004), crime (b=0.006), social cohesion (b=−0.002), greenness (b=−0.006), distance to hospitals, pharmacies and leisure services (b<0.001), O_3_ (b=0.001), winter temperature (b=0.010), as well as SD of summer (b=−0.001) and winter (b=0.008) temperature had non-zero coefficients (see α and λ in Table S8). After including also smoking, physical activity and BMI (sensitivity), leisure services for Horvath, grey space for Hannum, and crime, social cohesion, greenness, PM_10_, NO_2_, mean winter, and SD of summer and winter temperatures for DunedinPACE were selected (Table S9).

### Domains of exposure

Exploratory factor analysis identified a unidimensional structure within the social, built, retail, and health domains, with all retained exposures exhibiting adequate loadings (>0.25). Road traffic noise was excluded from the physico-chemical domain because of its low loading (0.11), and the resulting factor was interpreted as representing air pollution. The weather domain was not suitable for factor analysis (Kaiser–Meyer–Olkin=0.42) (Table S10; Figure S6).

In regression analysis, higher scores for social environment (b=0.423, 95%CI: 0.097, 0.749) and air pollution (b=0.386, 95%CI: 0.075, 0.697) were associated with PhenoAge. For DunedinPoAm, social environment (b=0.008, 95%CI: 0.003, 0.013), built environment (b=0.005, 95%CI: 0.001, 0.009) and air pollution (b=0.006, 95%CI: 0.001, 0.011), while for DunedinPACE social environment (b=0.018, 95%CI: 0.010, 0.025), built environment (b=0.012, 95%CI: 0.005, 0.018) and air pollution (b=0.017, 95%CI: 0.009, 0.024) were positively associated (Figure 2). Associations with DunedinPACE remained FDR-significant in the sensitivity analysis when adjusting for health behaviour (Table S11); multiple imputation did not change the findings (Figure S7). In mutually adjusted models including all domains, the associations between social environment and DunedinPACE remained robust, but air pollution, social environment and the health environment still showed nominal associations with both DunedinPoAm and DunedinPACE (Table S12).

**Figure 2.**
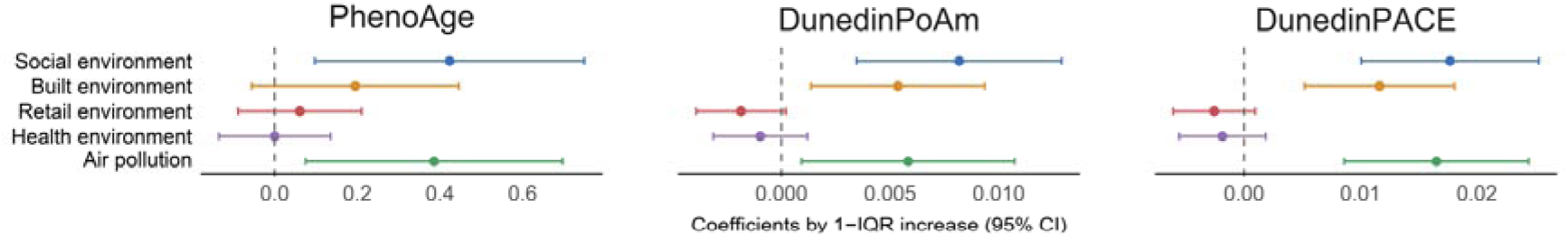
Associations of environmental domain scores with epigenetic clocks. Weighted linear regression models were fitted separately for each domain score–clock combination and estimates are expressed per interquartile range (IQR) increase in the domain score. Models were adjusted for age, sex, data collection wave, household income quartile, country of birth, housing tenure, partnership status, economic activity, government region, and technical variables summarised as a single component. Sample size was n = 3307.

### Exposure clusters

Cluster validation suggested either a two-cluster solution (silhouette) or a three-cluster solution (gap statistic and elbow method) (Figure S8). As the two-cluster version closely resembled the standard urban-rural classification available in the survey (i.e., 95% of participants in the urban cluster were also officially living in urban areas), we presented findings for this solution. Figure 3 shows average standardized environmental exposures in urban (n=2241; 73.2%) and rural (n=1066; 26.8%) clusters. In comparison to rural, urban residents had lower biological age as measured by Horvath’s clock (b=−0.385, 95%CI: −0.760, −0.011), but higher pace of ageing indicated by DunedinPoAm (b=0.008, 95%CI: 0.001, 0.014) and DunedinPACE (b=0.011, 95%CI: 0.000, 0.022) (Table 2). Sensitivity analysis showed attenuated, but persistent findings for Horvath’s clock and DunedinPoAm, and findings were robust to multiple imputation (Table S13). Three-cluster solution (i.e., urban, mixed, rural [reference]) (Figure S9) found comparable findings with membership in the urban cluster being associated with accelerated ageing, as reflected by PhenoAge, DunedinPoAm, and DunedinPACE, but not with Horvath’s clock (Table S14).

**Figure 3.**
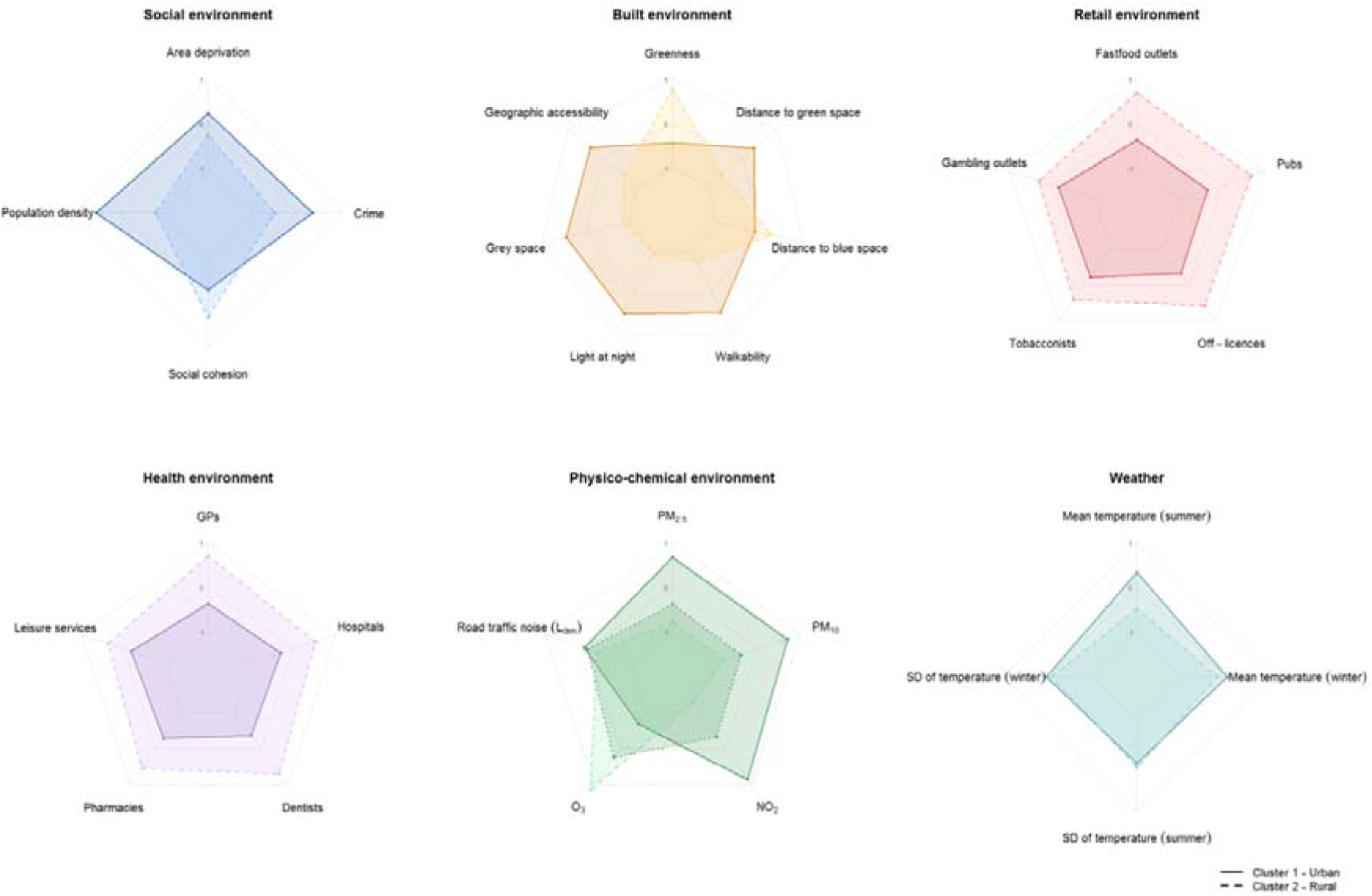
Weighted mean (95% CI) environmental exposures in urban and rural clusters. Values were scaled from their original unit. Sample size was n=1066 for the rural cluster and n=2241 for the urban cluster.

**Table 2.** Associations between urbanicity clusters and epigenetic clocks.

| Exposures | Model |  |  |
| --- | --- | --- | --- |
|  | b | 95% CI | p |
| Horvath |  |  |  |
| Urban (ref Rural) | -0.385 | -0.760, -0.011 | 0.044 |
| Hannum |  |  |  |
| Urban (ref Rural) | -0.100 | -0.386, 0.187 | 0.495 |
| PhenoAge |  |  |  |
| Urban (ref Rural) | -0.069 | -0.515, 0.378 | 0.764 |
| DunedinPoAm |  |  |  |
| Urban (ref Rural) | 0.008 | 0.001, 0.014 | 0.020 |
| DunedinPACE |  |  |  |
| Urban (ref Rural) | 0.011 | 0.000, 0.022 | 0.043 |

Weighted linear regression models were fitted separately for each epigenetic clock, with cluster membership (urban versus rural) as the exposure. Models were adjusted for age, sex, data collection wave, household income quartile, country of birth, housing tenure, partnership status, economic activity, government region, and technical variables summarised as a single component. Sample size was n = 3307.

## DISCUSSION

In this large, nationally representative study, leveraging four complementary analytical approaches, findings showed that adverse social, built and physico-chemical environments were associated with epigenetic ageing, with the strongest and most consistent signals observed for later-generation measures, particularly DunedinPACE. Across analyses, area deprivation, neighbourhood crime, lower social cohesion, lower greenness, higher PM_10_, NO_2_and O_3_ concentrations, and higher mean temperature emerged as most robust correlates of biological ageing. Associations with first-generation clocks were limited. Greater distance to leisure services was associated with higher biological age as measured by Horvath and PhenoAge. Finally, data-driven clustering identified urban and rural exposure profiles, with the former more robustly associated with faster pace of ageing.

The social environment was consistently associated with second- and third-generation clocks, particularly DunedinPACE, with area deprivation, neighbourhood crime and social cohesion emerging as key components after accounting for multicollinearity using elastic net models.

Area-level income deprivation, a marker of collective social and material resources, is a well-established social determinant of health linked to adverse health outcomes, independent of individual socioeconomic position. ^35^ A scoping review of nine studies reported associations between neighbourhood deprivation and epigenetic clocks, although findings varied by clock. ^13^ Consistent with large US-based studies, our results indicate that second- and third-generation clocks more sensitively capture biological signatures of social exposures than first-generation measures. ^36,37^ Evidence on non-economic social exposures remains limited. Living in high-crime neighbourhoods was associated with accelerated ageing across second- and third-generation clocks. While individual-level exposures, including interpersonal violence^38^ and childhood maltreatment, ^39^ have been linked to DNA methylation, the broader ecological effects of neighbourhood violence are less well characterized, with existing works largely focusing on children^40^, or related constructs such as neighbourhood disorder^37^. Given robust associations between area-level crime and population mental health independent of individual victimisation or deprivation^41^, chronic exposure to stressful neighbourhood conditions, such as crime hotspots, may plausibly shape DNA methylation profiles^42^. Greater neighbourhood social cohesion was associated with slower pace of ageing (DunedinPACE), in line with previous work^37^. Social cohesion has been linked to both mental^43^ and physical health^44^, with loneliness acting as a potential mediator^44^. In sensitivity analyses, adjustment for health behaviours substantially attenuated associations, suggesting partial mediation via smoking, physical activity, and adiposity, consistent with previous studies^37,45^.

Lower residential greenness, but not distance to nearest park, was associated with biological ageing, particularly for DunedinPoAm and DunedinPACE clocks (and for PhenoAge at the 1000m buffer). Although the evidence base remains limited, studies report that greater greenness, typically measured using NDVI, tree cover or proximity to green spaces, is associated with slower biological ageing. ^46–48^ However, prior studies primarily reported positive associations with GrimAge, which was not available in UKHLS. The only study to examine DunedinPACE reported null findings, ^48^ although it used a substantially larger buffer (5km) and differed in key sample characteristics. The choice of spatial buffer is a critical methodological consideration, as it should reflect population characteristics, relevant activity spaces and the outcome of interest^49^. In this study, we used 300m, 500m and 1000m buffers to capture variation in mobility-related local exposures, including older adults. Nonetheless, it is possible that larger buffers would yield different results.

A recent meta-analysis of 25 studies reported mixed evidence for associations between air pollution and epigenetic clocks, although particulate matter showed a trend toward accelerated biological ageing^12^. In our study, ambient air pollutants (PM_2.5_, PM_10_, and NO_2_) were positively associated with DunedinPACE, with somewhat weaker evidence of positive associations for DunedinPoAm and PhenoAge. Effect sizes for both the composite air pollution domain and individual pollutants in the ExWAS were 2.5-3.0-fold larger for DunedinPACE than for DunedinPoAm, suggesting greater sensitivity of DunedinPACE to air pollution–related variation in biological ageing. The only previous study using DunedinPACE similarly observed a positive association between NO_2_ exposure and pace of ageing among Black participants. ^50^ In contrast, ozone was associated with DunedinPACE in the opposite direction. As a secondary pollutant formed from NO_x_ via titration, it has typically higher concentrations in rural areas and is negatively associated with other pollutants (*r*=−0.87 with NO_2_ in our sample). This strong correlation may help explain the elastic net results, in which O_3_ was selected in the main model, whereas PM_10_ and NO_2_ were selected in sensitivity analyses. Air pollution may influence epigenetic alterations through multiple pathways, including oxidative stress driven by reactive oxygen species and altered expression of genes involved in inflammation, immune function, and nitric oxide pathways. ^51^

Higher average summer and winter temperatures were linked to accelerated pace of ageing (DunedinPoAm and DunedinPACE, respectively). Evidence indicates that short-, medium- and long-term exposure to elevated temperatures is linked to DNA methylation changes; for example, an Australian study identified 31 temperature-associated CpG sites implicated in disease pathways. ^52^ Higher annual temperatures have also been associated with age acceleration across multiple clocks, including Horvath, Hannum and PhenoAge. ^53^ Exposure to ambient temperature and heat stress, ^54^ as well as short-term temperature variability, particularly during warm season, have been linked to epigenetic clocks. ^55^ DunedinPACE has been associated with long-term exposure to ambient heat in a nationally representative US sample. ^56^ Mechanistically, heat may influence epigenetic ageing through disrupted thermoregulation, inflammation and oxidative stress, alongside temperature-related changes in physical activity. ^54–56^

Associations were stronger for second- and third-generation clocks than for first-generation measures. While first generation clocks were developed to predict chronological age, later clocks aimed to predict mortality and health outcomes by integrating clinical biomarkers across multiple physiological systems. Second- and third-generation clocks have shown improved predictive performance for health outcomes, such as mortality, multimorbidity, diseases (particularly respiratory and liver diseases) and depression^10,57^, and exhibit stronger associations with socioeconomic factors and health behaviours^57^. Only one environmental exposure—distance to leisure services—was significantly associated with a first-generation clock after FDR correction. Although distance to gambling outlets was also associated with Horvath and PhenoAge, this exposure was highly correlated with distance to leisure facilities (*r* = 0.83), and elastic net models only selected the latter, suggesting that the gambling-outlet association was not independent of correlated environmental features. Effect sizes were modest: a 1-IQR (∼4 km) increase in distance to leisure facilities was associated with 0.06 (Horvath) and 0.07 (PhenoAge) years of age acceleration, substantially smaller than estimates for other environmental exposures, albeit those were not significant. Physical activity is associated with slower biological ageing, ^58^ with structured aerobic and strength training shown to reduce epigenetic age acceleration. ^59^ While direct evidence using epigenetic clocks is limited, proximity to sport facilities is associated with higher frequency of exercise^60^ and more favourable health outcomes, including lower BMI, lower body fat percentage and smaller waist circumference. ^61^

Most existing studies rely on urban samples, potentially conflating environmental disadvantage with urbanicity. Evidence from representative samples remains limited and mixed. For example, one US study reported faster pace of ageing in rural areas using DunedinPACE, whereas previous analyses from the same cohort as ours found no difference using administrative urban-rural classification. ^62^ Our data-driven clustering identified an urban exposure profile associated with faster pace of ageing as measured by DunedinPoAm and DunedinPACE, which may reflect the co-occurrence of less favourable social, built and physico-chemical exposures in urban settings. By contrast, Horvath’s clock showed lower biological age in urban compared with rural clusters, although this pattern was observed only in the two-cluster solution. Although it is possible that rural disadvantage in access to services, including leisure facilities, contributes to this association, these findings should be interpreted cautiously because the clocks capture different aspects of biological ageing and may vary in their sensitivity to environmental conditions. Importantly, our data-driven clustering approach, based on observed environmental profiles rather than administrative classifications alone, provides a complementary way to characterise the environmental features that distinguish urban and rural settings and may contribute to differences in biological ageing.

This study provides a comprehensive assessment of the external exposome at residence in relation to epigenetic ageing, encompassing 30 exposures across multiple domains, including several environmental features that have rarely been examined, such as crime, retail, and health-service environments. Key strengths include the use of a nationally representative sample with survey weights, supporting generalisability to the White adult population of Great Britain; one of the largest samples in environmental epigenetics to date; and a multi-layered analytical framework that triangulated findings across four complementary approaches. The use of multiple spatial buffers further strengthened the robustness of the findings. Several limitations should be considered. First, exposure assessment relied on residential postcode-level geocoding, which, although relatively high resolution (∼0.13 km^2^), may introduce misclassification and remains subject to the uncertain geographic context and the modifiable areal unit problems. ^63^ Second, exposure-specific limitations remain (see EXPANSE^26^ and AHAH^27^ documentation), with some exposures, such as road traffic noise, ^64^ being less well characterised than others. Third, despite broad coverage of the environmental exposome, some dimension, particularly chemical exposures, were underrepresented. Fourth, DNA methylation data were available only for participants of White ethnicity in UKHLS, limiting generalisability to more diverse populations. Fifth, although all models were adjusted for a common set of confounders, different environmental exposures may operate through distinct confounding structures, and residual confounding is likely to remain. Finally, the cross-sectional design precludes causal inference. Although environmental conditions may contribute to biological ageing, reverse causation and residential selection remain plausible: individuals with poorer health, greater socioeconomic disadvantage or more advanced biological ageing may be more likely to reside in less advantaged neighbourhoods. ^65^ Future research should replicate these findings in longitudinal cohorts and evaluate the extent to which the reported associations reflect causal pathways.

## CONCLUSIONS

In this large, nationally representative study, we examined how the external residential exposome, spanning six domains and 30 exposures, relates to biological ageing. Adverse social, built and physico-chemical environments, including higher area-level deprivation, higher neighbourhood crime, lower social cohesion, reduced greenness, elevated air pollution and higher ambient temperatures, were associated with accelerated epigenetic ageing, particularly using second- and third-generation clocks. While limited associations were observed for first-generation measures (e.g., distance to leisure services with Horvath’s clock), newer clocks (e.g., DunedinPACE) demonstrated greater sensitivity in capturing the biological imprint of environmental exposures. Although the consistency of findings across multiple analytical approaches strengthens confidence in the observed associations, causal inference remains limited. Future research should investigate combined multidimensional environmental exposures in longitudinal studies, apply causal inference frameworks (e.g. quasi-experimental designs), and further elucidate biological pathways linking environmental exposures to healthy ageing.

## Data Availability

Data are available in a public, open access repository. This work uses Understanding Society: Waves 1-15, 2009-2024 and Harmonised BHPS: Waves 1-18, 1991-2009: Secure Access (SN: 6676), Understanding Society: Waves 1-14, 2009-2023 and Harmonised BHPS: Waves 1-18, 1991-2009 (SN: 6614) and Understanding Society: Waves 2-3 Nurse Health Assessment, 2010-2012 (SN: 7251). These datasets are available to researchers through the UK Data Service subject to registration and approval.

