## Supplementary Material for "Multidomain residential external exposome and epigenetic ageing in the UK general population"

**Online Supplement**

**Table S1:** Environmental and geographic variables used in the study

| **Domain** | **Exposure** | **Description** | **Resolution of input data** | | **Expressed as** | **Buffer** |
| --- | --- | --- | --- | --- | --- | --- |
|  |  |  | **Spatial** | **Temporal** |  |  |
| **Built environment** | Greenness | Normalized Difference Vegetation Index (NDVI) is an indicator of greenness (values range between -1 and 1) derived from MODIS satellite imagery; water features were removed using a high-resolution water mask layer (Joint Research Centre Global Surface Water). Original 250 x 250 m resolution data was resampled to 100 x 100m resolution.^1^ | 100 × 100m | 2010 | index (from -1 to 1) | 300m |
|  | Distance to green space | Euclidean distance to nearest publicly accessible green land cover types using the CORINE map. Green cover types included were urban parks and sport and leisure facilities, agricultural areas and forest and semi-natural areas. Distance is measured from the pixel centroid.^1^ | 100 × 100m | 2012 | meters | NA |
|  | Distance to blue space | Euclidean distance to nearest inland water feature or seashore. The EU-Hydro dataset was used to characterize the indicator. Distance is measured from the pixel centroid.^1^ | 100 × 100m | 2013 | meters | NA |
|  | Walkability | Composite index of walkability (sum of z-scores) in a geographic unit, computed from a range of indicators correlated with walking:^1^  *Walkability = z(population density) + 0.5*z(street connectivity) + 0.5*z(pedestrian street density) – z(slope) – z(distance to green spaces) + z(food stores density) + z(NDVI) + z(public transport station density)* | 100 × 100m | 2020 | index | 300m |
|  | Light at night | Computed using Defense Meteorological Satellite Program for the years 2010.^1^ | 100 × 100m | 2010 | radiance | 300m |
|  | Grey space | Imperviousness is characterized by the substitution of the original (semi-) natural land cover or water surface with an artificial, often impenetrable or sealed cover, and expressed as the percentage of soil sealing per area unit. It was derived using the imperviousness degree layer from the Copernicus Land Surface Monitoring Service for the year of 2010.^1^ | 100 × 100m | 2012 | percentage | 300m |
|  | Geographic accessibility | Geographic access to services, such as GP surgery, supermarket, school, or post office. Ranks of access domains from the English^2^, Welsh^3^ and Scottish^4^ indices of multiple deprivation were divided into 10 equal groups within each country, with higher deciles expressing better access. *NOTE*: there are significant differences in how the indicators in different countries were derived (e.g. distance using walking, bus journey, road distance). | E/W: Lower Super Output Area (LSOA)  S: Data zone | E: 2010  W: 2011  S: 2012 | Deciles; 1 (lowest access) to 10 (highest access) | NA |
| **Physico-chemical environment** | Air pollution | Geographically and temporally weighted regression was used to compute monthly concentrations by regressing observations from routine monitoring stations on several spatial predictor variables, such as chemical transport model estimates, satellite-derived data, meteorological data, and land-use and road variables. Models showed satisfactory performance, explaining 71-82% of the variance in PM_2.5_ levels, 50-68% of the variance in PM_10_ levels, 61-69% of the variance in NO_2_ levels, 45-67% of the variance in O_3_ levels as given by 5-fold cross-validated R^2^ values.^5^ Area-weighted average values of air pollution were derived using 500 meter buffer around residential postcodes.  Pollutants:   a) PM_2.5_ (fine particles with a diameter of <2.5µm) b) PM_10_ (fine particles with a diameter of <10µm) c) NO_2_ (nitrogen dioxide) d) O_3_ (ozone) | 100 × 100m | 12-month exposures before interview | µg m^-3^ | 300m around postcode centroids |
|  | Road traffic noise (L_den_) | 24 hours weighted road traffic noise exposure along major roads (>3m vehicle annually) and within agglomerations of >100.000 people. Noise levels were estimated on a 10m grid at the receptor height of 4 meters for the year of 2012, following the Environmental Noise Directive (Directive 2002/49/EC). Noise classes were separately estimated across England,^6^ Wales^7^ and Scotland.^8^ We harmonised country-specific noise classes into 3 common classes. | NA | 2012 | Noise classes in decibel (dB):  ≤54.9 dB  55.00-59.9 dB  60.00-64.9 dB  ≥65 dB | NA |
| **Weather** | Temperature | Ambient temperature was modelled using random forest approach, with measurements obtained from global, regional and local ground monitoring networks. Predictors included: Land Surface Temperature, CORINE Land-Cover classes, impervious surface density, NDVI, latitude and longitude, population density, and meteorological variables from the 5^th^ generation ECMWF atmospheric reanalysis of the global climate. Models explained on average 96% of the variability in minimum, maximum and mean daily temperature measurements, as given by 5-fold cross-validated R^2^ values.^5^   a) Mean temperature, warm season (i.e. computed from monthly average temperature between April-September) b) Standard deviation of temperature, warm season (i.e. computed from monthly average temperature between April-September) c) Mean temperature, cold season (i.e. computed from monthly average temperature between March-October) d) Standard deviation of temperature, cold season (i.e. computed from monthly average temperature between March-October) | 1km × 1km | 12-month exposures before interview | Celsius degree (ºC) | NA |
| **Social environment** | Area deprivation | Proportion of the population in a geographic area with indication of low income-based benefits (e.g. Income Support, Job Seeker Allowance, Child Tax Credits). Ranks of income domains from the English^2^, Welsh^3^ and Scottish^4^ indices of multiple deprivation were divided into 10 equal groups within each country, with higher deciles expressing higher deprivation. *NOTE*: there are differences in how the indicators in different countries were derived. | E/W: LSOA  S: Data zone | E: 2010  W: 2011  S: 2012 | Deciles; 1 (lowest deprivation) to 10 (highest deprivation) | NA |
|  | Crime | Ranks of crime domains from the English^2^, Welsh^3^ and Scottish^4^ indices of multiple deprivation were divided into 10 equal groups within each country, with higher deciles expressing higher crime. *NOTE*: there are significant differences in how the indicators in different countries were derived. | E/W: LSOA  S: Data zone | E: 2010  W: 2011  S: 2012 | Deciles; 1 (lowest crime) to 10 (highest crime) | NA |
|  | Social cohesion (aggregated) | A shortened version of Buckner's Neighbourhood Cohesion scale^9^ was used in Understanding Society to measure attraction to the neighbourhood, neighbouring, and sense of community using 8 questions (i.e., 1. Belong to neighbourhood; 2. Local friends mean a lot; 3. Advice obtainable locally; 4. Can borrow things from neighbours; 5. Willing to improve neighbourhood; 6. Plan to stay in neighbourhood; 7. Being similar to others in neighbourhood; 8. Talk regularly to neighbours) with 5 response options on a Likert scale (1-lowest cohesion; 5-highest cohesion). Item responses were averaged for each respondent ranging from 1 to 5. Because the scale was not assessed in every wave, we used responses from all available participants in the waves temporally closest to the outcome measurement (i.e. UKHLS wave 1 [2009/2010; n=38555]; UKHLS wave 3 [2011/2012; n=40560], BHPS wave 18 [2008; n=13166]), and subsequently calculated average values for each LSOA/Data Zone. | E/W: LSOA  S: Data zone | 2008,  2009/2010,  2011/2012, | Index (1 to 5) | NA |
|  | Population density | Population density was derived from the Global Human Settlement Layer, a degree of population density was calculated on a continuous scale which higher scores indicating more urban surface. Focal mean within a 1500m square was applied to create 100 × 100m surface.^1^ | 100 × 100m | 2015 | index | NA |
| **Retail environment** | Fast food outlets | Nearest fast food outlets (Fish & Chips Shops, Indian Takeaway, Pizza Takeaway, Sandwich Delivery Service, Take Away Food Shops) based on road network distance from each postcode centroid, aggregated into higher geographies. Taken from the Access to Healthy Assets and Hazards index (version 1)^10^ | E/W: LSOA  S: Data zone | 2016 | kilometres | NA |
|  | Pubs | Nearest pub/bar/nightclub (Night Clubs, Bars, Public Houses & Inns) based on road network distance from each postcode centroid, aggregated into higher geographies. Taken from the Access to Healthy Assets and Hazards index (version 1)^10^ | E/W: LSOA  S: Data zone | 2016 | kilometres | NA |
|  | Off-licences | Nearest off licence based on road network distance from each postcode centroid, aggregated into higher geographies. Taken from the Access to Healthy Assets and Hazards index (version 1)^10^ | E/W: LSOA  S: Data zone | 2016 | kilometres | NA |
|  | Tobacconists | Nearest tobacconist based on road network distance from each postcode centroid, aggregated into higher geographies. Taken from the Access to Healthy Assets and Hazards index (version 1)^10^ | E/W: LSOA  S: Data zone | 2016 | kilometres | NA |
|  | Gambling outlets | Nearest gambling outlet (Casino Clubs, Bookmakers) based on road network distance from each postcode centroid, aggregated into higher geographies. Taken from the Access to Healthy Assets and Hazards index (version 1)^10^ | E/W: LSOA  S: Data zone | 2016 | kilometres | NA |
| **Health environment** | GPs | Nearest GP practice based on road network distance from each postcode centroid, aggregated into higher geographies. Taken from the Access to Healthy Assets and Hazards index (version 1)^10^ | E/W: LSOA  S: Data zone | 2016 | kilometres | NA |
|  | Hospitals | Nearest A&E hospitals based on road network distance from each postcode centroid, aggregated into higher geographies. Taken from the Access to Healthy Assets and Hazards index (version 1)^10^ | E/W: LSOA  S: Data zone | 2016 | kilometres | NA |
|  | Dentists | Nearest dentist based on road network distance from each postcode centroid, aggregated into higher geographies. Taken from the Access to Healthy Assets and Hazards index (version 1)^10^ | E/W: LSOA  S: Data zone | 2016 | kilometres | NA |
|  | Pharmacies | Nearest pharmacy based on road network distance from each postcode centroid, aggregated into higher geographies. Taken from the Access to Healthy Assets and Hazards index (version 1)^10^ | E/W: LSOA  S: Data zone | 2016 | kilometres | NA |
|  | Leisure services | Nearest leisure service based on road network distance from each postcode centroid, aggregated into higher geographies. Taken from the Access to Healthy Assets and Hazards index (version 1)^10^ | E/W: LSOA  S: Data zone | 2016 | kilometres | NA |
| **Geography** | Regions | Eleven regions across Great Britain (i.e. North East, North West, Yorkshire and the Humber, East Midlands, West Midlands, East of England, London, South East, South West, Wales, and Scotland) based on the NUT1 (Nomenclature of territorial units for statistics, Level 1) classification (i.e. Government Office Regions) | Regions | 2010 | NA | NA |

**Figure S1**: Schematic directed acyclic graph representing exposures, outcomes and confounders. Dashed lines indicate confounders, which were only included in the sensitivity analysis as they might be on the causal path between exposures and outcomes.

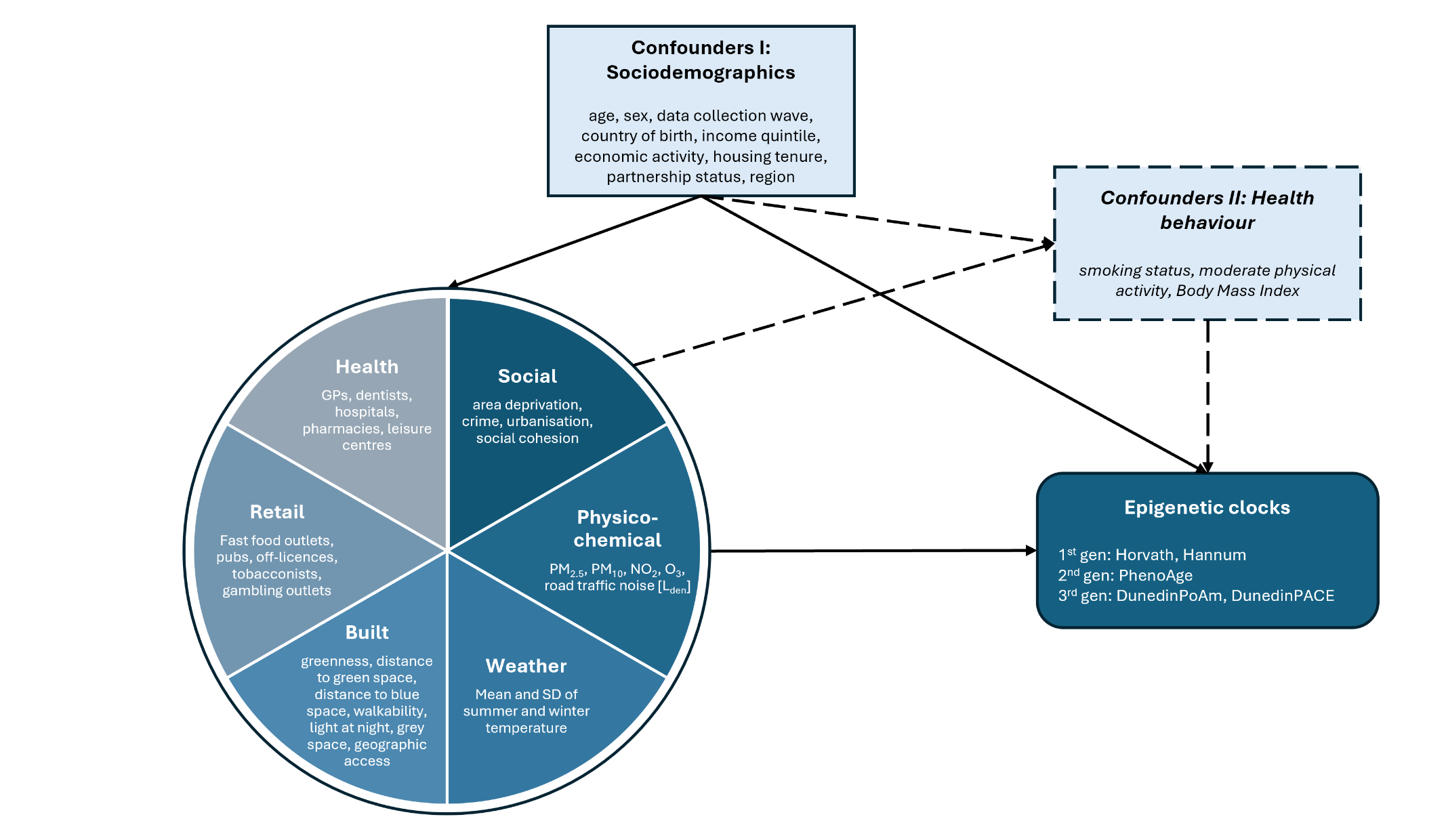

**Table S2**: Mean and interquartile range of units of environmental exposures.

| **Exposure** |  | | **Exposure** |  | |
| --- | --- | --- | --- | --- | --- |
|  | Mean | Interquartile range (IQR) |  | Mean | Interquartile range (IQR) |
| Social environment | | | Health environment | | |
| Area deprivation | 5.05 | 4.00 | GPs | 1.81 | 1.29 |
| Crime | 5.05 | 4.00 | Hospitals | 12.65 | 11.40 |
| Social cohesion | 3.64 | 0.54 | Dentists | 2.06 | 1.48 |
| Population density | 28.60 | 26.04 | Pharmacies | 1.65 | 0.95 |
| Built environment | | | Leisure services | 5.68 | 4.16 |
| Greenness | 0.54 | 0.11 | Physico-chemical environment | | |
| Distance to green space | 336.40 | 347.00 | Road traffic noise | NA |  |
| Distance to blue space | 1832.96 | 1560.09 | PM_2.5_ | 11.82 | 3.20 |
| Walkability | 13.12 | 4.00 | PM_10_ | 17.98 | 4.11 |
| Light at night | 48.78 | 20.00 | NO_2_ | 21.91 | 10.00 |
| Grey space | 34.27 | 32.00 | O_3_ | 58.09 | 5.73 |
| Geographic accessibility | 5.07 | 4.00 | Temperature | | |
| Retail environment | | | Mean temp (summer) | 15.48 | 1.29 |
| Fast food outlets | 2.90 | 2.53 | Mean temp (winter) | 4.07 | 2.02 |
| Pubs | 2.17 | 1.57 | SD of temp (summer) | 0.82 | 0.32 |
| Off-licences | 6.09 | 6.20 | SD of temp (winter) | 2.02 | 2.27 |
| Tobacconists | 6.75 | 5.68 |  |  |  |
| Gambling outlets | 3.21 | 2.41 |  |  |  |

Values are presented on their original scales as weighted means and IQRs. Area deprivation, crime and geographic accessibility are expressed as deciles, with higher values indicating greater deprivation, crime and accessibility. Social cohesion is a composite scale, with higher values indicating greater cohesion; population density is expressed as population per hectare; greenness is measured using the Normalised Difference Vegetation Index (NDVI); and distance to green and blue space is expressed in metres. Walkability is a composite index, light at night is measured in radiance, and grey space represents the percentage of built-up land cover. Distances to fast-food outlets, pubs, off-licences, tobacconists, gambling outlets, GPs, hospitals, dentists, pharmacies and leisure services are expressed in kilometres. Road traffic noise is categorised into decibel classes (1: ≤54.9 dB; 2: 55.0–59.9 dB; 3: 60.0–64.9 dB; 4: ≥65.0 dB). PM_2.5_, PM_10_, NO_2_, O_3_ are expressed in µg m^-3^, and temperature is expressed in degrees Celsius. The sample size was n=3613.

**Figure S2**: Flowchart for sample selection

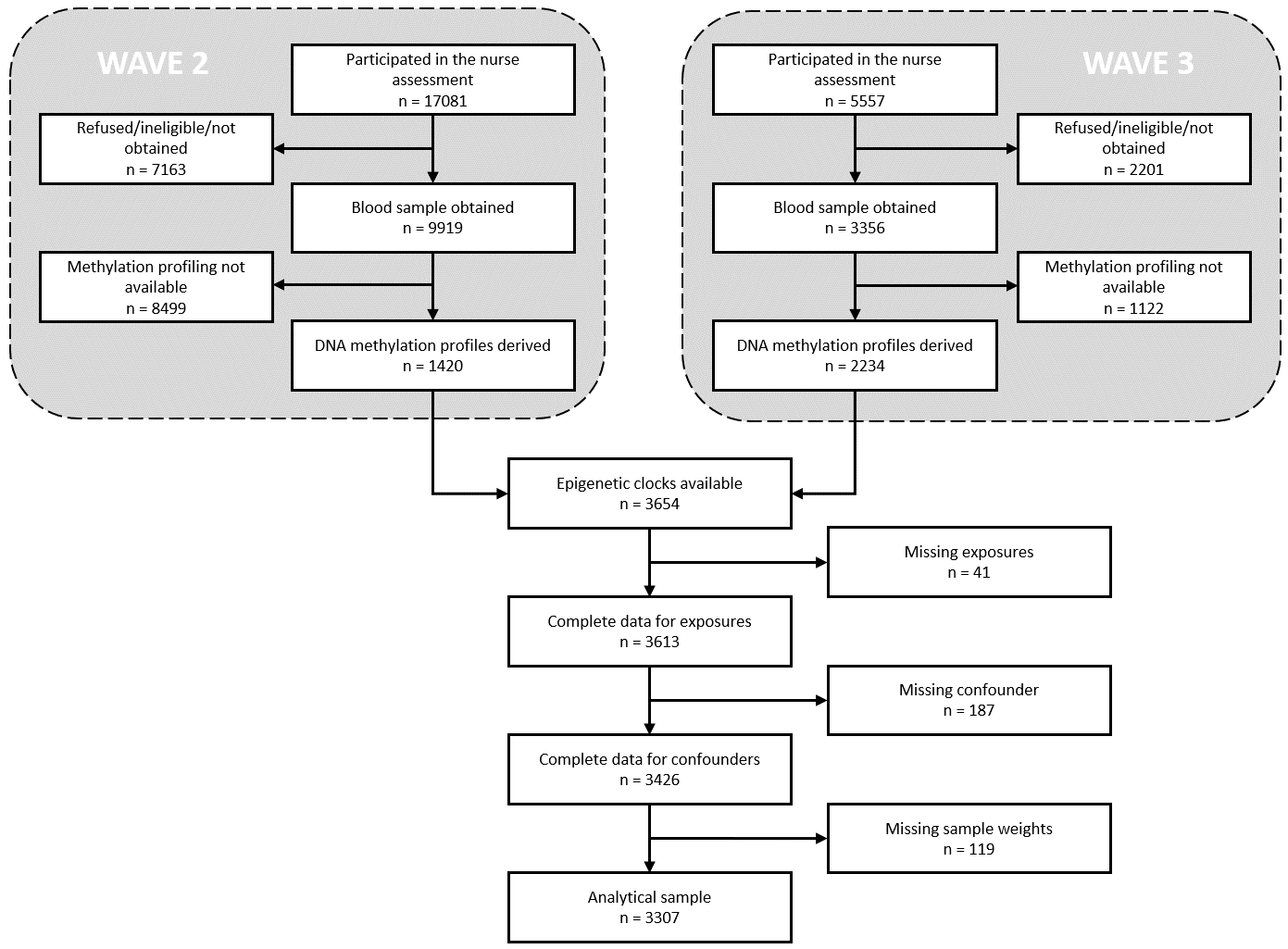

**Figure S3:** Correlation between environmental exposures. Pearson’s coefficients are presented in the analytical sample of n=3307.

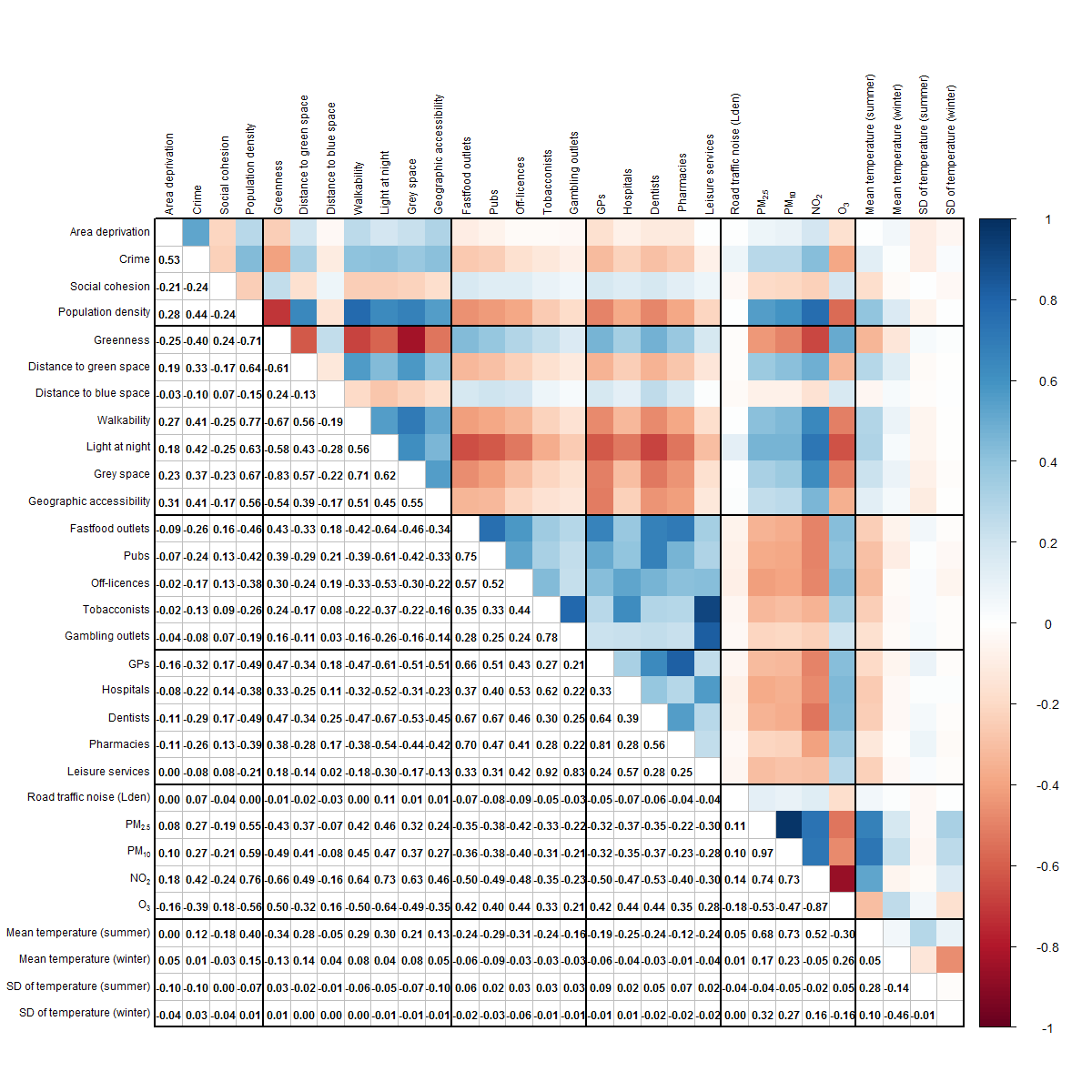

**Table S3**: Single exposure models for Horvath’s clock, main and sensitivity analysis 1.

| **Exposures** | **Main model** | | | | **Sensitivity 1** | | | |
| --- | --- | --- | --- | --- | --- | --- | --- | --- |
|  | b | 95% CI | p | p_FDR_ | b | 95% CI | p | p_FDR_ |
| Area deprivation | 0.044 | -0.220, 0.308 | 0.744 | 0.930 | 0.020 | -0.243, 0.282 | 0.883 | 0.995 |
| Crime | 0.001 | -0.244, 0.247 | 0.991 | 0.991 | -0.002 | -0.247, 0.243 | 0.985 | 0.995 |
| Social cohesion | -0.010 | -0.196, 0.177 | 0.919 | 0.991 | -0.003 | -0.188, 0.181 | 0.973 | 0.995 |
| Population density | -0.079 | -0.275, 0.117 | 0.430 | 0.912 | -0.091 | -0.287, 0.105 | 0.362 | 0.836 |
| Greenness | 0.101 | -0.098, 0.299 | 0.321 | 0.912 | 0.106 | -0.095, 0.306 | 0.302 | 0.836 |
| Distance to green space | 0.004 | -0.169, 0.176 | 0.968 | 0.991 | 0.009 | -0.159, 0.176 | 0.918 | 0.995 |
| Distance to blue space | 0.065 | -0.145, 0.274 | 0.545 | 0.912 | 0.044 | -0.162, 0.251 | 0.673 | 0.962 |
| Walkability | -0.006 | -0.226, 0.215 | 0.959 | 0.991 | -0.001 | -0.219, 0.218 | 0.995 | 0.995 |
| Light at night | -0.043 | -0.271, 0.186 | 0.714 | 0.930 | -0.042 | -0.272, 0.188 | 0.722 | 0.985 |
| Grey space | -0.256 | -0.50, -0.012 | 0.040 | 0.238 | -0.260 | -0.504, -0.016 | 0.037 | 0.220 |
| Geographic accessibility | -0.089 | -0.363, 0.185 | 0.524 | 0.912 | -0.129 | -0.403, 0.145 | 0.356 | 0.836 |
| Fast food outlets | 0.018 | -0.107, 0.142 | 0.778 | 0.934 | 0.014 | -0.113, 0.141 | 0.828 | 0.995 |
| Pubs | 0.030 | -0.075, 0.134 | 0.578 | 0.912 | 0.023 | -0.083, 0.129 | 0.671 | 0.962 |
| Off-licences | 0.103 | -0.048, 0.255 | 0.180 | 0.675 | 0.096 | -0.055, 0.246 | 0.211 | 0.764 |
| Tobacconists | 0.072 | 0.020, 0.124 | 0.007 | 0.071 | 0.077 | 0.016, 0.138 | 0.013 | 0.121 |
| Gambling outlets | **0.035** | **0.022, 0.048** | **<0.001** | **<0.001** | **0.041** | **0.028, 0.055** | **<0.001** | **<0.001** |
| GPs | 0.070 | -0.039, 0.178 | 0.208 | 0.692 | 0.067 | -0.042, 0.177 | 0.229 | 0.764 |
| Hospitals | 0.068 | -0.093, 0.229 | 0.406 | 0.912 | 0.052 | -0.109, 0.213 | 0.526 | 0.962 |
| Dentists | 0.006 | -0.126, 0.139 | 0.929 | 0.991 | 0.001 | -0.134, 0.135 | 0.993 | 0.995 |
| Pharmacies | 0.041 | -0.012, 0.094 | 0.133 | 0.649 | 0.042 | -0.012, 0.095 | 0.127 | 0.563 |
| Leisure services | **0.060** | **0.030, 0.091** | **<0.001** | **0.001** | **0.065** | **0.032, 0.098** | **<0.001** | **0.002** |
| Road traffic noise | -0.066 | -0.432, 0.299 | 0.721 | 0.930 | -0.041 | -0.405, 0.323 | 0.825 | 0.995 |
| PM_2.5_ | 0.143 | -0.157, 0.442 | 0.350 | 0.912 | 0.143 | -0.155, 0.441 | 0.348 | 0.836 |
| PM_10_ | 0.071 | -0.229, 0.371 | 0.642 | 0.930 | 0.068 | -0.230, 0.366 | 0.657 | 0.962 |
| NO_2_ | -0.078 | -0.350, 0.195 | 0.575 | 0.912 | -0.085 | -0.358, 0.189 | 0.544 | 0.962 |
| O_3_ | -0.087 | -0.380, 0.205 | 0.558 | 0.912 | -0.073 | -0.366, 0.219 | 0.623 | 0.962 |
| Mean temp (summer) | -0.359 | -0.665, -0.053 | 0.022 | 0.162 | -0.376 | -0.682, -0.070 | 0.016 | 0.121 |
| Mean temp (winter) | 0.055 | -0.197, 0.307 | 0.667 | 0.930 | 0.072 | -0.176, 0.320 | 0.571 | 0.962 |
| SD of temp (summer) | -0.067 | -0.246, 0.112 | 0.463 | 0.912 | -0.075 | -0.252, 0.102 | 0.405 | 0.868 |
| SD of temp (winter) | 0.220 | -0.081, 0.520 | 0.152 | 0.649 | 0.227 | -0.068, 0.522 | 0.131 | 0.563 |

Weighted linear regression models were fitted separately for each exposure-outcome combination, expressed per interquartile range (IQR) increase. The main models adjusted for age, sex, wave of data collection, income quartile, born in the UK, housing tenure, partnership status, employment status, government regions, and technical variables (as a single component). In the sensitivity analysis, we additionally adjusted for smoking status, moderate physical activity, and body mass index. Variance inflation factors were <10. Sample size was n=3307.

**Table S4**: Single exposure models for Hannum’s clock, main and sensitivity analysis 1.

| **Exposures** | **Main model** | | | | **Sensitivity 1** | | | |
| --- | --- | --- | --- | --- | --- | --- | --- | --- |
|  | b | 95% CI | p | p_FDR_ | b | 95% CI | p | p_FDR_ |
| Area deprivation | -0.013 | -0.238, 0.213 | 0.913 | 0.961 | -0.049 | -0.277, 0.179 | 0.674 | 0.990 |
| Crime | 0.095 | -0.091, 0.282 | 0.315 | 0.961 | 0.071 | -0.118, 0.260 | 0.461 | 0.990 |
| Social cohesion | -0.015 | -0.159, 0.129 | 0.839 | 0.961 | -0.009 | -0.151, 0.134 | 0.906 | 0.990 |
| Population density | 0.021 | -0.145, 0.186 | 0.806 | 0.961 | 0.013 | -0.154, 0.179 | 0.882 | 0.990 |
| Greenness | 0.023 | -0.143, 0.189 | 0.787 | 0.961 | 0.036 | -0.133, 0.206 | 0.675 | 0.990 |
| Distance to green space | 0.078 | -0.069, 0.225 | 0.298 | 0.961 | 0.076 | -0.074, 0.226 | 0.319 | 0.990 |
| Distance to blue space | -0.054 | -0.185, 0.077 | 0.420 | 0.961 | -0.049 | -0.180, 0.081 | 0.459 | 0.990 |
| Walkability | 0.015 | -0.162, 0.193 | 0.866 | 0.961 | 0.011 | -0.169, 0.190 | 0.906 | 0.990 |
| Light at night | 0.020 | -0.153, 0.194 | 0.820 | 0.961 | 0.015 | -0.161, 0.190 | 0.870 | 0.990 |
| Grey space | -0.079 | -0.273, 0.114 | 0.421 | 0.961 | -0.088 | -0.283, 0.108 | 0.380 | 0.990 |
| Geographic accessibility | 0.037 | -0.189, 0.263 | 0.745 | 0.961 | 0.009 | -0.219, 0.238 | 0.936 | 0.990 |
| Fast food outlets | 0.017 | -0.063, 0.097 | 0.682 | 0.961 | 0.015 | -0.065, 0.096 | 0.712 | 0.990 |
| Pubs | 0.017 | -0.052, 0.085 | 0.632 | 0.961 | 0.016 | -0.054, 0.086 | 0.655 | 0.990 |
| Off-licences | 0.027 | -0.067, 0.120 | 0.578 | 0.961 | 0.023 | -0.070, 0.117 | 0.622 | 0.990 |
| Tobacconists | -0.002 | -0.031, 0.028 | 0.912 | 0.961 | -0.006 | -0.037, 0.026 | 0.730 | 0.990 |
| Gambling outlets | 0.001 | -0.006, 0.009 | 0.738 | 0.961 | 0.000 | -0.009, 0.009 | 0.961 | 0.990 |
| GPs | -0.029 | -0.118, 0.060 | 0.526 | 0.961 | -0.026 | -0.117, 0.065 | 0.574 | 0.990 |
| Hospitals | -0.015 | -0.120, 0.091 | 0.784 | 0.961 | -0.028 | -0.134, 0.079 | 0.610 | 0.990 |
| Dentists | -0.043 | -0.124, 0.039 | 0.306 | 0.961 | -0.043 | -0.125, 0.039 | 0.303 | 0.990 |
| Pharmacies | 0.002 | -0.036, 0.040 | 0.929 | 0.961 | 0.004 | -0.034, 0.042 | 0.849 | 0.990 |
| Leisure services | 0.008 | -0.010, 0.027 | 0.385 | 0.961 | 0.006 | -0.015, 0.026 | 0.581 | 0.990 |
| Road traffic noise | 0.112 | -0.185, 0.410 | 0.459 | 0.961 | 0.126 | -0.165, 0.416 | 0.396 | 0.990 |
| PM_2.5_ | 0.243 | 0.016, 0.471 | 0.036 | 0.569 | 0.237 | 0.008, 0.467 | 0.042 | 0.668 |
| PM_10_ | 0.251 | 0.014, 0.489 | 0.038 | 0.569 | 0.245 | 0.006, 0.484 | 0.045 | 0.668 |
| NO_2_ | 0.004 | -0.203, 0.211 | 0.971 | 0.971 | -0.001 | -0.209, 0.207 | 0.990 | 0.990 |
| O_3_ | -0.034 | -0.259, 0.192 | 0.770 | 0.961 | -0.036 | -0.263, 0.191 | 0.756 | 0.990 |
| Mean temp (summer) | -0.075 | -0.324, 0.174 | 0.553 | 0.961 | -0.080 | -0.334, 0.173 | 0.534 | 0.990 |
| Mean temp (winter) | 0.100 | -0.108, 0.309 | 0.346 | 0.961 | 0.092 | -0.118, 0.302 | 0.390 | 0.990 |
| SD of temp (summer) | -0.095 | -0.235, 0.045 | 0.184 | 0.961 | -0.098 | -0.238, 0.043 | 0.172 | 0.990 |
| SD of temp (winter) | 0.190 | -0.048, 0.429 | 0.118 | 0.961 | 0.188 | -0.052, 0.429 | 0.125 | 0.990 |

Weighted linear regression models were fitted separately for each exposure-outcome combination, expressed per interquartile range (IQR) increase. The main models adjusted for age, sex, wave of data collection, income quartile, born in the UK, housing tenure, partnership status, employment status, government regions, and technical variables (as a single component). In the sensitivity analysis, we additionally adjusted for smoking status, moderate physical activity, and body mass index. Variance inflation factors were <10. Sample size was n=3307.

**Table S5**: Single exposure models for PhenoAge, main and sensitivity analysis 1.

| **Exposures** | **Main model** | | | | **Sensitivity 1** | | | |
| --- | --- | --- | --- | --- | --- | --- | --- | --- |
|  | b | 95% CI | p | p_FDR_ | b | 95% CI | p | p_FDR_ |
| Area deprivation | 0.353 | 0.022, 0.683 | 0.036 | 0.140 | 0.183 | -0.135, 0.502 | 0.258 | 0.581 |
| Crime | 0.399 | 0.098, 0.701 | 0.010 | 0.096 | 0.276 | -0.016, 0.569 | 0.064 | 0.240 |
| Social cohesion | -0.043 | -0.294, 0.208 | 0.738 | 0.763 | -0.022 | -0.264, 0.221 | 0.861 | 0.891 |
| Population density | 0.221 | -0.009, 0.450 | 0.060 | 0.156 | 0.163 | -0.059, 0.385 | 0.149 | 0.407 |
| Greenness | -0.112 | -0.346, 0.122 | 0.348 | 0.615 | -0.021 | -0.258, 0.215 | 0.860 | 0.891 |
| Distance to green space | 0.223 | 0.013, 0.434 | 0.037 | 0.140 | 0.213 | 0.012, 0.414 | 0.038 | 0.189 |
| Distance to blue space | -0.122 | -0.319, 0.075 | 0.225 | 0.421 | -0.088 | -0.278, 0.102 | 0.364 | 0.683 |
| Walkability | 0.191 | -0.057, 0.438 | 0.130 | 0.301 | 0.149 | -0.092, 0.390 | 0.225 | 0.562 |
| Light at night | 0.178 | -0.077, 0.434 | 0.170 | 0.365 | 0.120 | -0.128, 0.367 | 0.344 | 0.683 |
| Grey space | 0.022 | -0.262, 0.306 | 0.880 | 0.880 | -0.064 | -0.345, 0.217 | 0.655 | 0.854 |
| Geographic accessibility | 0.112 | -0.219, 0.444 | 0.506 | 0.695 | -0.020 | -0.345, 0.305 | 0.905 | 0.905 |
| Fast food outlets | -0.036 | -0.168, 0.095 | 0.589 | 0.695 | -0.022 | -0.152, 0.109 | 0.742 | 0.890 |
| Pubs | -0.038 | -0.140, 0.065 | 0.471 | 0.695 | -0.029 | -0.137, 0.079 | 0.597 | 0.852 |
| Off-licences | 0.070 | -0.096, 0.236 | 0.407 | 0.678 | 0.067 | -0.093, 0.228 | 0.411 | 0.724 |
| Tobacconists | 0.067 | 0.002, 0.133 | 0.044 | 0.148 | 0.075 | 0.006, 0.144 | 0.034 | 0.189 |
| Gambling outlets | **0.036** | **0.021, 0.050** | **<0.001** | **<0.001** | **0.043** | **0.029, 0.057** | **<0.001** | **<0.001** |
| GPs | -0.031 | -0.162, 0.100 | 0.643 | 0.715 | -0.014 | -0.142, 0.113 | 0.824 | 0.891 |
| Hospitals | 0.050 | -0.118, 0.217 | 0.559 | 0.695 | 0.016 | -0.140, 0.173 | 0.836 | 0.891 |
| Dentists | -0.076 | -0.197, 0.045 | 0.217 | 0.421 | -0.066 | -0.184, 0.052 | 0.271 | 0.581 |
| Pharmacies | -0.029 | -0.119, 0.062 | 0.534 | 0.695 | -0.019 | -0.104, 0.065 | 0.651 | 0.854 |
| Leisure services | **0.066** | **0.035, 0.097** | **<0.001** | **<0.001** | **0.070** | **0.037, 0.104** | **<0.001** | **0.001** |
| Road traffic noise | 0.080 | -0.344, 0.504 | 0.710 | 0.761 | 0.143 | -0.282, 0.567 | 0.510 | 0.765 |
| PM_2.5_ | 0.424 | 0.077, 0.772 | 0.017 | 0.108 | 0.359 | 0.020, 0.698 | 0.038 | 0.189 |
| PM_10_ | 0.425 | 0.073, 0.777 | 0.018 | 0.108 | 0.363 | 0.023, 0.703 | 0.037 | 0.189 |
| NO_2_ | 0.280 | -0.014, 0.575 | 0.062 | 0.156 | 0.224 | -0.068, 0.515 | 0.132 | 0.397 |
| O_3_ | -0.370 | -0.691, -0.048 | 0.024 | 0.122 | -0.326 | -0.647, -0.006 | 0.046 | 0.196 |
| Mean temp (summer) | 0.133 | -0.245, 0.510 | 0.490 | 0.695 | 0.067 | -0.293, 0.426 | 0.717 | 0.890 |
| Mean temp (winter) | -0.098 | -0.404, 0.207 | 0.528 | 0.695 | -0.120 | -0.420, 0.181 | 0.435 | 0.724 |
| SD of temp (summer) | -0.057 | -0.269, 0.156 | 0.603 | 0.695 | -0.069 | -0.272, 0.133 | 0.501 | 0.765 |
| SD of temp (winter) | 0.347 | -0.015, 0.709 | 0.061 | 0.156 | 0.319 | -0.034, 0.672 | 0.076 | 0.254 |

Weighted linear regression models were fitted separately for each exposure-outcome combination, expressed per interquartile range (IQR) increase. The main models adjusted for age, sex, wave of data collection, income quartile, born in the UK, housing tenure, partnership status, employment status, government regions, and technical variables (as a single component). In the sensitivity analysis, we additionally adjusted for smoking status, moderate physical activity, and body mass index. Variance inflation factors were <10. Sample size was n=3307.

**Table S6**: Single exposure models for DunedinPoAm, main and sensitivity analysis 1.

| **Exposures** | **Main model** | | | | **Sensitivity 1** | | | |
| --- | --- | --- | --- | --- | --- | --- | --- | --- |
|  | b | 95% CI | p | p_FDR_ | b | 95% CI | p | p_FDR_ |
| Area deprivation | **0.007** | **0.003, 0.012** | **0.002** | **0.012** | 0.003 | -0.001, 0.006 | 0.174 | 0.379 |
| Crime | **0.007** | **0.003, 0.011** | **0.001** | **0.012** | 0.003 | 0.000, 0.006 | 0.080 | 0.379 |
| Social cohesion | -0.001 | -0.004, 0.002 | 0.439 | 0.598 | 0.000 | -0.003, 0.002 | 0.745 | 0.894 |
| Population density | 0.004 | 0.000, 0.008 | 0.059 | 0.146 | 0.002 | -0.001, 0.006 | 0.254 | 0.477 |
| Greenness | **-0.006** | **-0.009, -0.002** | **0.001** | **0.012** | -0.002 | -0.005, 0.001 | 0.115 | 0.379 |
| Distance to green space | 0.002 | -0.001, 0.005 | 0.222 | 0.370 | 0.002 | -0.001, 0.004 | 0.169 | 0.379 |
| Distance to blue space | -0.001 | -0.004, 0.001 | 0.364 | 0.546 | 0.001 | -0.001, 0.003 | 0.356 | 0.619 |
| Walkability | 0.004 | -0.001, 0.008 | 0.095 | 0.191 | 0.002 | -0.002, 0.005 | 0.399 | 0.630 |
| Light at night | 0.005 | 0.001, 0.009 | 0.022 | 0.084 | 0.002 | -0.001, 0.005 | 0.135 | 0.379 |
| Grey space | 0.005 | 0.001, 0.010 | 0.022 | 0.084 | 0.002 | -0.002, 0.005 | 0.371 | 0.619 |
| Geographic accessibility | 0.004 | -0.001, 0.009 | 0.122 | 0.228 | 0.000 | -0.004, 0.004 | 0.966 | 0.966 |
| Fast food outlets | -0.002 | -0.004, 0.000 | 0.068 | 0.146 | -0.001 | -0.003, 0.000 | 0.103 | 0.379 |
| Pubs | -0.002 | -0.003, 0.000 | 0.038 | 0.115 | -0.001 | -0.002, 0.000 | 0.158 | 0.379 |
| Off-licences | -0.002 | -0.004, 0.001 | 0.144 | 0.253 | -0.001 | -0.003, 0.001 | 0.190 | 0.379 |
| Tobacconists | 0.000 | -0.001, 0.001 | 0.389 | 0.556 | 0.000 | -0.001, 0.001 | 0.962 | 0.966 |
| Gambling outlets | 0.000 | 0.000, 0.000 | 0.614 | 0.697 | 0.000 | 0.000, 0.000 | 0.432 | 0.643 |
| GPs | -0.001 | -0.003, 0.002 | 0.627 | 0.697 | 0.000 | -0.002, 0.001 | 0.880 | 0.966 |
| Hospitals | -0.001 | -0.003, 0.002 | 0.462 | 0.603 | -0.001 | -0.003, 0.001 | 0.188 | 0.379 |
| Dentists | -0.002 | -0.004, 0.000 | 0.046 | 0.126 | -0.001 | -0.003, 0.000 | 0.054 | 0.379 |
| Pharmacies | 0.000 | -0.001, 0.001 | 0.960 | 0.973 | 0.000 | -0.001, 0.001 | 0.804 | 0.928 |
| Leisure services | 0.000 | -0.001, 0.001 | 0.973 | 0.973 | 0.000 | 0.000, 0.001 | 0.450 | 0.643 |
| Road traffic noise | -0.003 | -0.009, 0.003 | 0.360 | 0.546 | 0.000 | -0.005, 0.005 | 0.950 | 0.966 |
| PM_2.5_ | 0.006 | 0.001, 0.011 | 0.021 | 0.084 | 0.003 | 0.000, 0.007 | 0.071 | 0.379 |
| PM_10_ | 0.007 | 0.001, 0.012 | 0.016 | 0.084 | 0.004 | 0.000, 0.008 | 0.037 | 0.379 |
| NO_2_ | 0.005 | 0.000, 0.010 | 0.038 | 0.115 | 0.003 | -0.001, 0.006 | 0.152 | 0.379 |
| O_3_ | -0.005 | -0.010, 0.000 | 0.065 | 0.146 | -0.003 | -0.007, 0.001 | 0.116 | 0.379 |
| Mean temp (summer) | **0.009** | **0.003, 0.014** | **0.001** | **0.012** | **0.006** | **0.002, 0.010** | **0.001** | **0.043** |
| Mean temp (winter) | 0.000 | -0.004, 0.005 | 0.862 | 0.924 | -0.001 | -0.004, 0.002 | 0.506 | 0.690 |
| SD of temp (summer) | 0.001 | -0.002, 0.004 | 0.582 | 0.697 | 0.001 | -0.002, 0.003 | 0.538 | 0.699 |
| SD of temp (winter) | 0.002 | -0.004, 0.007 | 0.581 | 0.697 | 0.001 | -0.003, 0.005 | 0.559 | 0.699 |

Weighted linear regression models were fitted separately for each exposure-outcome combination, expressed per interquartile range (IQR) increase. The main models adjusted for age, sex, wave of data collection, income quartile, born in the UK, housing tenure, partnership status, employment status, government regions, and technical variables (as a single component). In the sensitivity analysis, we additionally adjusted for smoking status, moderate physical activity, and body mass index. Variance inflation factors were <10. Sample size was n=3307.

**Table S7**: Single exposure models for DunedinPACE, main and sensitivity analysis 1.

| **Exposures** | **Main model** | | | | **Sensitivity 1** | | | |
| --- | --- | --- | --- | --- | --- | --- | --- | --- |
|  | b | 95% CI | p | p_FDR_ | b | 95% CI | p | p_FDR_ |
| Area deprivation | **0.013** | **0.005, 0.020** | **0.001** | **0.004** | 0.005 | -0.002, 0.011 | 0.189 | 0.314 |
| Crime | **0.011** | **0.004, 0.019** | **0.004** | **0.011** | 0.005 | -0.002, 0.011 | 0.141 | 0.250 |
| Social cohesion | **-0.009** | **-0.014, -0.003** | **0.001** | **0.006** | **-0.008** | **-0.012, -0.003** | **0.002** | **0.015** |
| Population density | **0.010** | **0.004, 0.017** | **0.002** | **0.007** | 0.007 | 0.001, 0.013 | 0.017 | 0.057 |
| Greenness | **-0.012** | **-0.018, -0.006** | **<0.001** | **<0.001** | **-0.007** | **-0.013, -0.002** | **0.006** | **0.028** |
| Distance to green space | 0.005 | 0.000, 0.010 | 0.071 | 0.120 | 0.004 | 0.000, 0.009 | 0.063 | 0.156 |
| Distance to blue space | 0.000 | -0.005, 0.004 | 0.893 | 0.924 | 0.002 | -0.002, 0.006 | 0.417 | 0.547 |
| Walkability | **0.009** | **0.001, 0.016** | **0.018** | **0.045** | 0.006 | 0.000, 0.013 | 0.06 | 0.156 |
| Light at night | 0.008 | 0.001, 0.015 | 0.025 | 0.057 | 0.004 | -0.001, 0.01 | 0.136 | 0.250 |
| Grey space | **0.013** | **0.006, 0.021** | **0.001** | **0.004** | 0.009 | 0.002, 0.015 | 0.012 | 0.050 |
| Geographic accessibility | 0.009 | 0.001, 0.018 | 0.034 | 0.073 | 0.003 | -0.005, 0.011 | 0.422 | 0.547 |
| Fast food outlets | -0.003 | -0.007, 0.001 | 0.159 | 0.238 | -0.002 | -0.005, 0.001 | 0.238 | 0.375 |
| Pubs | -0.003 | -0.006, 0.000 | 0.071 | 0.120 | -0.002 | -0.005, 0.000 | 0.068 | 0.156 |
| Off-licences | 0.000 | -0.004, 0.004 | 0.837 | 0.924 | 0.000 | -0.004, 0.003 | 0.917 | 0.946 |
| Tobacconists | 0.000 | -0.002, 0.001 | 0.601 | 0.751 | 0.000 | -0.001, 0.001 | 0.58 | 0.696 |
| Gambling outlets | 0.000 | -0.001, 0.000 | 0.190 | 0.272 | 0.000 | 0.000, 0.000 | 0.864 | 0.926 |
| GPs | -0.002 | -0.007, 0.002 | 0.259 | 0.353 | -0.002 | -0.005, 0.002 | 0.36 | 0.514 |
| Hospitals | -0.001 | -0.005, 0.004 | 0.755 | 0.906 | -0.002 | -0.005, 0.002 | 0.297 | 0.445 |
| Dentists | -0.003 | -0.006, 0.000 | 0.072 | 0.120 | -0.002 | -0.004, 0.000 | 0.105 | 0.211 |
| Pharmacies | 0.000 | -0.002, 0.002 | 0.937 | 0.937 | 0.000 | -0.002, 0.002 | 0.792 | 0.880 |
| Leisure services | 0.000 | -0.001, 0.001 | 0.880 | 0.924 | 0.000 | 0.000, 0.001 | 0.437 | 0.547 |
| Road traffic noise | -0.001 | -0.012, 0.010 | 0.839 | 0.924 | 0.002 | -0.009, 0.013 | 0.701 | 0.809 |
| PM_2.5_ | **0.018** | **0.010, 0.027** | **<0.001** | **<0.001** | **0.014** | **0.007, 0.022** | **<0.001** | **0.001** |
| PM_10_ | **0.018** | **0.010, 0.027** | **<0.001** | **<0.001** | **0.015** | **0.008, 0.023** | **<0.001** | **0.001** |
| NO_2_ | **0.013** | **0.006, 0.021** | **0.001** | **0.004** | **0.010** | **0.003, 0.017** | **0.003** | **0.026** |
| O_3_ | **-0.014** | **-0.023, -0.005** | **0.001** | **0.006** | **-0.011** | **-0.019, -0.003** | **0.004** | **0.027** |
| Mean temp (summer) | 0.004 | -0.006, 0.013 | 0.446 | 0.582 | 0.000 | -0.007, 0.008 | 0.946 | 0.946 |
| Mean temp (winter) | **0.010** | **0.002, 0.018** | **0.016** | **0.045** | 0.009 | 0.002, 0.016 | 0.015 | 0.057 |
| SD of temp (summer) | -0.005 | -0.010, 0.001 | 0.118 | 0.187 | -0.005 | -0.010, 0.000 | 0.052 | 0.155 |
| SD of temp (winter) | 0.009 | -0.001, 0.019 | 0.072 | 0.12 | 0.007 | -0.001, 0.016 | 0.095 | 0.203 |

Weighted linear regression models were fitted separately for each exposure-outcome combination, expressed per interquartile range (IQR) increase. The main models adjusted for age, sex, wave of data collection, income quartile, born in the UK, housing tenure, partnership status, employment status, government regions, and technical variables (as a single component). In the sensitivity analysis, we additionally adjusted for smoking status, moderate physical activity, and body mass index. Variance inflation factors were <10. Sample size was n=3307.

**Figure S4:** Association between environmental exposures and epigenetic clocks in the main analysis and after multiple imputation. Weighted linear regression models were fitted separately for each exposure, expressed per interquartile range (IQR) increase. Models adjusted for age, sex, wave of data collection, income quartile, born in the UK, housing tenure, partnership status, employment status, government regions, and technical variables (as a single component). In the sensitivity analysis, missing data was imputed using multiple imputation by chained equations for 10 datasets and pooled based on Rubin’s rule. Sample size is n=3307 for the main, and n=3613 for the sensitivity analysis.

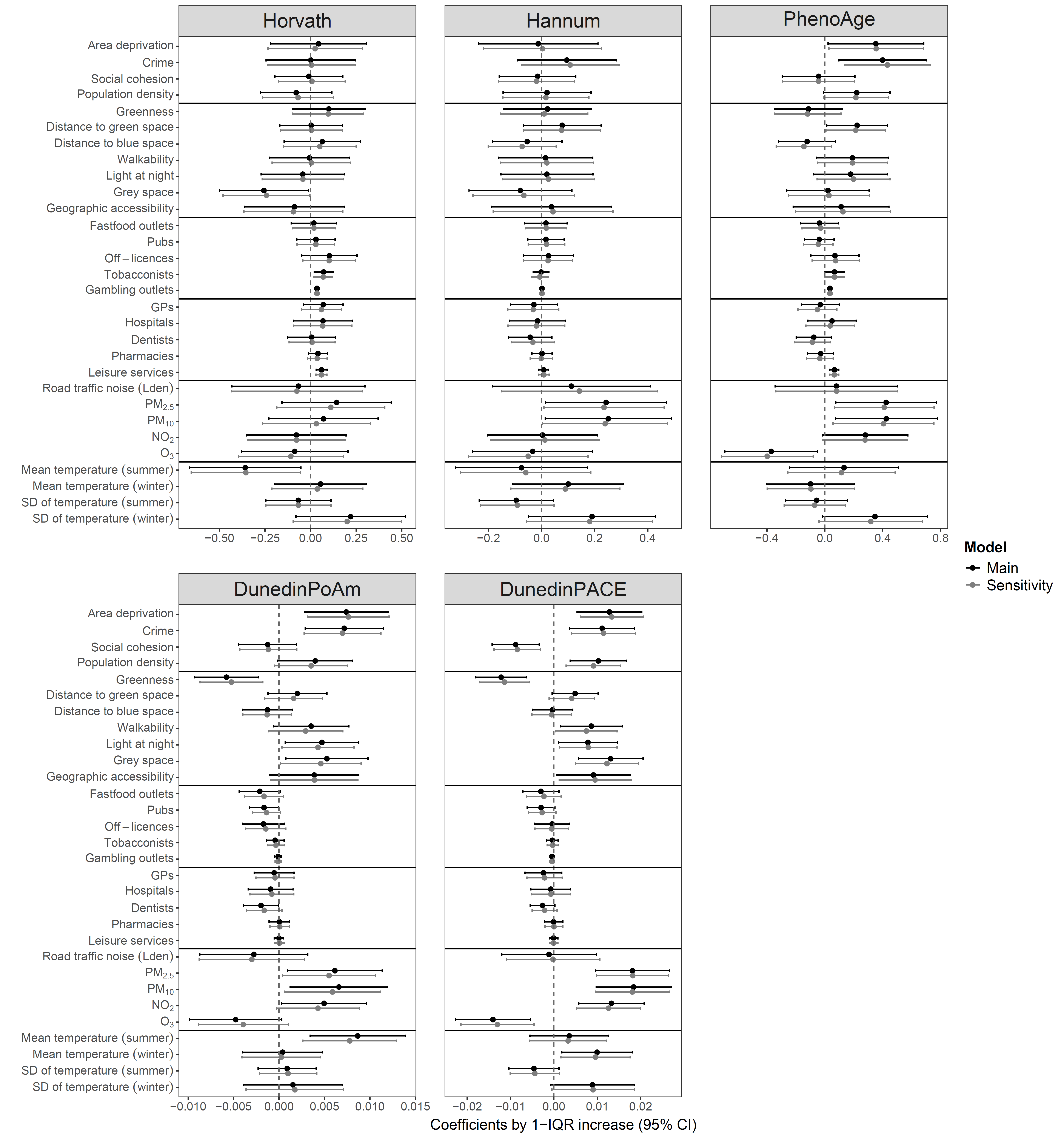

**Figure S5:** Single exposure models with 300m, 500m, 1000m buffers. Weighted linear regression models were fitted separately for each exposure, expressed per interquartile range (IQR) increase. The main models adjusted for age, sex, wave of data collection, income quartile, born in the UK, housing tenure, partnership status, employment status, government regions, and technical variables (as a single component). Sample size was n=3307.

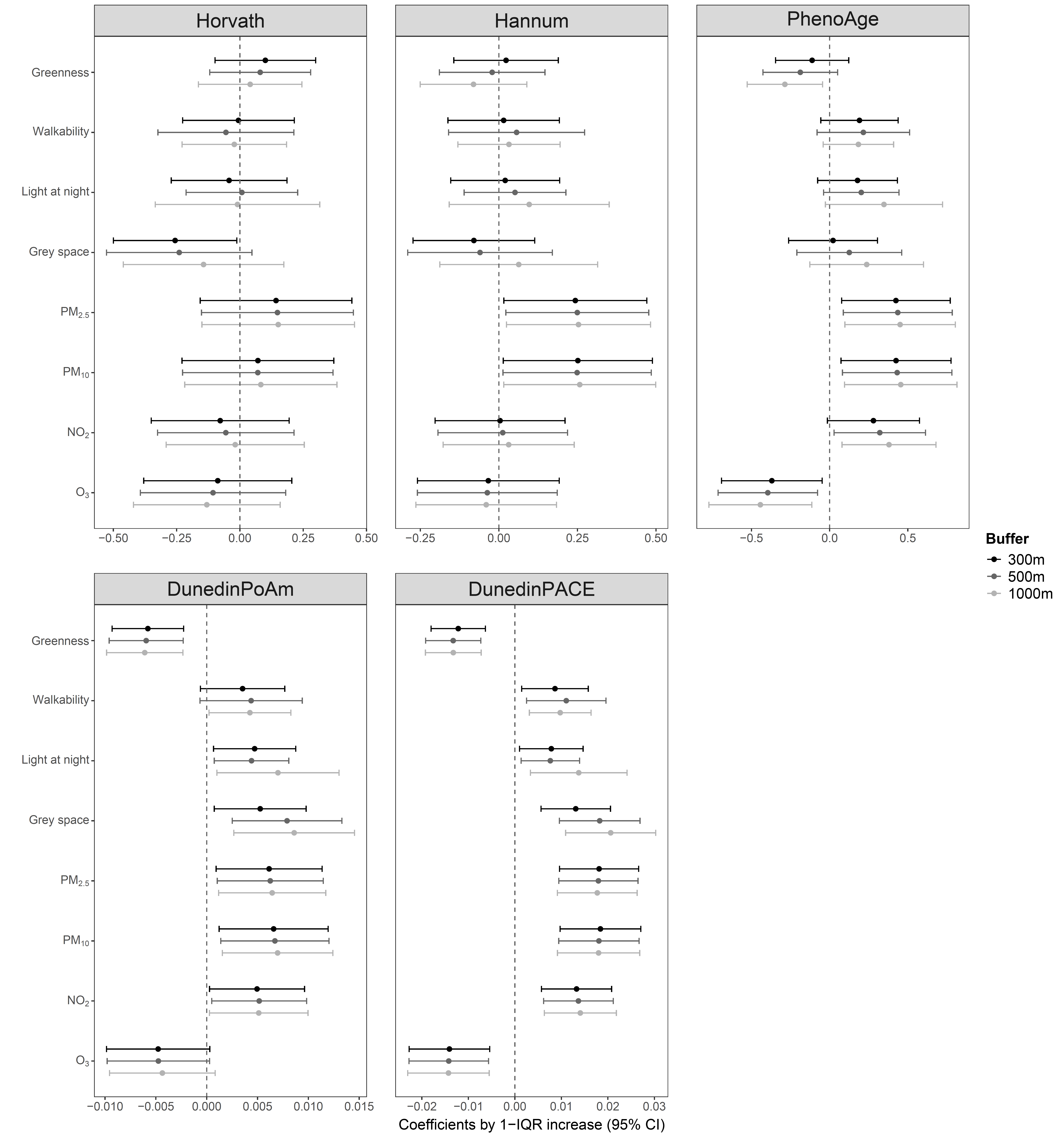

**Table S8**: Variable selection with elastic net regression, main analysis.

| **Exposures** | **Horvath** | **Hannum** | **PhenoAge** | **DunedinPoAm** | **DunedinPACE** |
| --- | --- | --- | --- | --- | --- |
| Area deprivation | ns | ns | ns | 0.003 | 0.004 |
| Crime | ns | <0.001 | 0.179 | 0.003 | 0.006 |
| Social cohesion | ns | ns | ns | ns | -0.002 |
| Population density | ns | ns | ns | ns | ns |
| Greenness | ns | ns | ns | -0.002 | -0.006 |
| Distance to green space | ns | ns | ns | ns | ns |
| Distance to blue space | ns | ns | ns | ns | ns |
| Walkability | ns | ns | ns | ns | ns |
| Light at night | ns | ns | ns | ns | ns |
| Grey space | ns | ns | ns | ns | ns |
| Geographic accessibility | ns | ns | ns | ns | ns |
| Fast food outlets | ns | ns | ns | ns | ns |
| Pubs | ns | ns | ns | ns | ns |
| Off-licences | ns | ns | ns | ns | ns |
| Tobacconists | ns | ns | ns | ns | ns |
| Gambling outlets | ns | ns | ns | ns | ns |
| GPs | ns | ns | ns | ns | ns |
| Hospitals | ns | ns | ns | ns | <0.001 |
| Dentists | ns | ns | ns | ns | ns |
| Pharmacies | ns | ns | ns | ns | <0.001 |
| Leisure services | <0.001 | ns | 0.001 | ns | <0.001 |
| Road traffic noise | ns | ns | ns | ns | ns |
| PM_2.5_ | ns | ns | ns | ns | ns |
| PM_10_ | ns | ns | ns | ns | ns |
| NO_2_ | ns | ns | ns | ns | ns |
| O_3_ | ns | ns | ns | ns | 0.001 |
| Mean temp (summer) | ns | ns | ns | ns | ns |
| Mean temp (winter) | ns | ns | ns | ns | 0.010 |
| SD of temp (summer) | ns | ns | ns | ns | -0.001 |
| SD of temp (winter) | ns | ns | ns | ns | 0.008 |
| *α* | 0.1 | 0.1 | 1.0 | 0.8 | 1.0 |
| *λ* | 0.888 | 0.440 | 0.099 | 0.001 | 0.001 |

The main models adjusted for age, sex, wave of data collection, income quartile, born in the UK, housing tenure, partnership status, employment status, government regions, and technical variables (as a single component). Sample size was n=3307. Abbreviations: ns=not selected.

**Table 9**: Variable selection with elastic net regression, sensitivity analysis 1.

| **Exposures** | **Horvath** | **Hannum** | **PhenoAge** | **DunedinPoAm** | **DunedinPACE** |
| --- | --- | --- | --- | --- | --- |
| Area deprivation | ns | ns | ns | ns | ns |
| Crime | ns | ns | ns | ns | 0.001 |
| Social cohesion | ns | ns | ns | ns | -0.001 |
| Population density | ns | ns | ns | ns | ns |
| Greenness | ns | ns | ns | ns | <0.001 |
| Distance to green space | ns | ns | ns | ns | ns |
| Distance to blue space | ns | ns | ns | ns | ns |
| Walkability | ns | ns | ns | ns | ns |
| Light at night | ns | ns | ns | ns | ns |
| Grey space | ns | <0.001 | ns | ns | ns |
| Geographic accessibility | ns | ns | ns | ns | ns |
| Fast food outlets | ns | ns | ns | ns | ns |
| Pubs | ns | ns | ns | ns | ns |
| Off-licences | ns | ns | ns | ns | ns |
| Tobacconists | ns | ns | ns | ns | ns |
| Gambling outlets | ns | ns | ns | ns | ns |
| GPs | ns | ns | ns | ns | ns |
| Hospitals | ns | ns | ns | ns | ns |
| Dentists | ns | ns | ns | ns | ns |
| Pharmacies | ns | ns | ns | ns | ns |
| Leisure services | <0.001 | ns | ns | ns | ns |
| Road traffic noise | ns | ns | ns | ns | ns |
| PM_2.5_ | ns | ns | ns | ns | ns |
| PM_10_ | ns | ns | ns | ns | 0.002 |
| NO_2_ | ns | ns | ns | ns | 0.002 |
| O_3_ | ns | ns | ns | ns | ns |
| Mean temp (summer) | ns | ns | ns | ns | ns |
| Mean temp (winter) | ns | ns | ns | ns | 0.003 |
| SD of temp (summer) | ns | ns | ns | ns | <0.001 |
| SD of temp (winter) | ns | ns | ns | ns | <0.001 |
| *α* | 0.1 | 0.1 | 0.1 | 0.1 | 0.1 |
| *λ* | 0.789 | 0.361 | 0.972 | 0.009 | 0.011 |

The sensitivity analysis adjusted for age, sex, wave of data collection, income quartile, born in the UK, housing tenure, partnership status, employment status, government regions, technical variables (as a single component), smoking status, moderate physical activity, and body mass index. Sample size was n=3307. Abbreviations: ns=not selected.

**Table S10**: Exploratory factor analysis on environmental exposures.

| **Domain** | **Environmental feature** | **Factor loading** |
| --- | --- | --- |
| Social environment  Kaiser-Meyer-Olkin value=0.66  Bartlett’s test of sphericity: χ^2^=2105.8 (p<0.001)  Variance explained: 36% | Income deprivation | 0.62 |
|  | Crime | 0.84 |
|  | Social cohesion | -0.31 |
|  | Population density | 0.52 |
| Built environment  Kaiser-Meyer-Olkin value=0.89  Bartlett’s test of sphericity: χ ^2^=11653.2 (p<0.001)  Variance explained: 51% | Greenness | -0.90 |
|  | Distance to green space | 0.66 |
|  | Distance to blue space | -0.26 |
|  | Walkability | 0.78 |
|  | Light at night | 0.68 |
|  | Grey space | 0.91 |
|  | Geographic accessibility | 0.61 |
| Retail environment  Kaiser-Meyer-Olkin value=0.65  Bartlett’s test of sphericity: χ ^2^= 8276.9 (p<0.001)  Variance explained: 45% | Fast-food outlets | 0.88 |
|  | Pubs | 0.83 |
|  | Off-licences | 0.65 |
|  | Tobacconists | 0.47 |
|  | Gambling outlets | 0.37 |
| Health environment  Kaiser-Meyer-Olkin value=0.70  Bartlett’s test of sphericity: χ ^2^= 7222.6 (p<0.001)  Variance explained: 46% | GPs | 0.94 |
|  | Hospitals | 0.37 |
|  | Dentists | 0.68 |
|  | Pharmacies | 0.86 |
|  | Leisure services | 0.29 |
| Physico-chemical environment  Kaiser-Meyer-Olkin value=0.61  Bartlett’s test of sphericity: χ ^2^=17920.8 (p<0.001)  Variance explained: 56%^a^ | Road traffic noise | *0.11* |
|  | PM_2.5_ | 1.00 |
|  | PM_10_ | 0.97 |
|  | NO_2_ | 0.74 |
|  | O_3_ | -0.53 |
| Weather  Kaiser-Meyer-Olkin value=0.42  Bartlett’s test of sphericity: χ ^2^=1289.3 (p<0.001)  Variance explained: 31% | Mean temperature (summer) | *0.05* |
|  | Mean temperature (winter) | 1.00 |
|  | SD of temperature (summer) | *-0.14* |
|  | SD of temperature (winter) | -0.46 |

Grey highlighted factor leadings are below the threshold of <0.25, thus dropped from the principal component. Sample size was n=3307.

^a^Increased to 69% after dropping road traffic noise.

**Figure S6**: Scree plots indicating eigenvalues of principal components (PC) and factor analysis (FA).

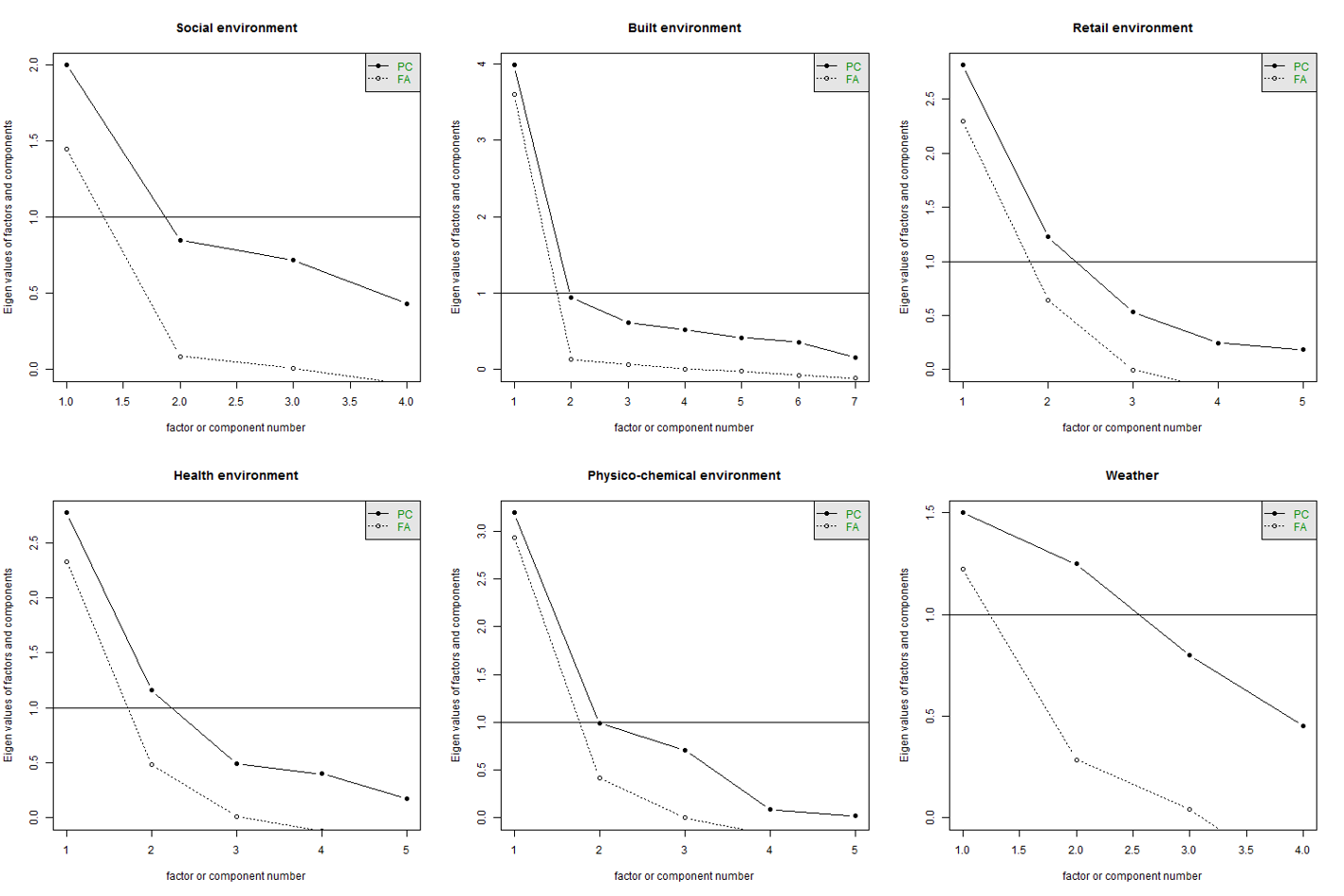

**Table S11**: Associations between exposure domains and epigenetic clocks, main and sensitivity analysis 1.

| **Exposures** | **Main model** | | | | **Sensitivity 1** | | | |
| --- | --- | --- | --- | --- | --- | --- | --- | --- |
|  | b | 95% CI | p | p_FDR_ | b | 95% CI | p | p_FDR_ |
| Horvath clock |  |  |  |  |  |  |  |  |
| Social environment | -0.014 | -0.279, 0.251 | 0.919 | 0.919 | -0.032 | -0.298, 0.233 | 0.810 | 0.810 |
| Built environment | -0.104 | -0.328, 0.120 | 0.361 | 0.602 | -0.107 | -0.332, 0.118 | 0.351 | 0.586 |
| Retail environment | 0.109 | -0.018, 0.236 | 0.094 | 0.341 | 0.112 | -0.026, 0.250 | 0.110 | 0.395 |
| Health environment | 0.095 | -0.030, 0.220 | 0.136 | 0.341 | 0.092 | -0.036, 0.220 | 0.158 | 0.395 |
| Air pollution | 0.049 | -0.231, 0.330 | 0.731 | 0.914 | 0.043 | -0.238, 0.323 | 0.766 | 0.810 |
| Hannum clock |  |  |  |  |  |  |  |  |
| Social environment | 0.054 | -0.155, 0.263 | 0.611 | 0.833 | 0.024 | -0.188, 0.236 | 0.824 | 0.957 |
| Built environment | 0.017 | -0.159, 0.192 | 0.853 | 0.853 | 0.005 | -0.174, 0.184 | 0.957 | 0.957 |
| Retail environment | 0.017 | -0.059, 0.092 | 0.666 | 0.833 | 0.012 | -0.065, 0.089 | 0.764 | 0.957 |
| Health environment | -0.020 | -0.104, 0.065 | 0.648 | 0.833 | -0.022 | -0.108, 0.064 | 0.617 | 0.957 |
| Air pollution | 0.129 | -0.085, 0.343 | 0.237 | 0.833 | 0.125 | -0.090, 0.341 | 0.255 | 0.957 |
| PhenoAge |  |  |  |  |  |  |  |  |
| Social environment | **0.423** | **0.097, 0.749** | **0.011** | **0.037** | 0.273 | -0.046, 0.592 | 0.093 | 0.233 |
| Built environment | 0.195 | -0.055, 0.445 | 0.127 | 0.211 | 0.111 | -0.136, 0.358 | 0.379 | 0.474 |
| Retail environment | 0.061 | -0.089, 0.211 | 0.424 | 0.530 | 0.078 | -0.076, 0.232 | 0.320 | 0.474 |
| Health environment | 0.000 | -0.135, 0.135 | 0.996 | 0.996 | 0.010 | -0.123, 0.143 | 0.886 | 0.886 |
| Air pollution | **0.386** | **0.075, 0.697** | **0.015** | **0.037** | 0.327 | 0.021, 0.632 | 0.036 | 0.180 |
| DunedinPoAm |  |  |  |  |  |  |  |  |
| Social environment | **0.008** | **0.003, 0.013** | **0.001** | **0.003** | 0.003 | 0.000, 0.007 | 0.076 | 0.190 |
| Built environment | **0.005** | **0.001, 0.009** | **0.009** | **0.022** | 0.002 | -0.001, 0.005 | 0.216 | 0.360 |
| Retail environment | -0.002 | -0.004, 0.000 | 0.077 | 0.096 | -0.001 | -0.002, 0.001 | 0.298 | 0.372 |
| Health environment | -0.001 | -0.003, 0.001 | 0.380 | 0.380 | -0.001 | -0.002, 0.001 | 0.504 | 0.504 |
| Air pollution | **0.006** | **0.001, 0.011** | **0.020** | **0.033** | 0.003 | 0.000, 0.007 | 0.062 | 0.190 |
| DunedinPACE |  |  |  |  |  |  |  |  |
| Social environment | **0.018** | **0.010, 0.025** | **<0.001** | **<0.001** | **0.010** | **0.003, 0.017** | **0.004** | **0.009** |
| Built environment | **0.012** | **0.005, 0.018** | **<0.001** | **0.001** | **0.007** | **0.002, 0.013** | **0.013** | **0.022** |
| Retail environment | -0.003 | -0.006, 0.001 | 0.153 | 0.192 | -0.001 | -0.004, 0.002 | 0.400 | 0.433 |
| Health environment | -0.002 | -0.006, 0.002 | 0.328 | 0.328 | -0.001 | -0.004, 0.002 | 0.433 | 0.433 |
| Air pollution | **0.017** | **0.009, 0.024** | **<0.001** | **<0.001** | **0.013** | **0.006, 0.020** | **<0.001** | **0.001** |

Weighted linear regression models were fitted separately for each exposure-outcome combination, expressed per interquartile range (IQR) increase. The main models adjusted for age, sex, wave of data collection, income quartile, born in the UK, housing tenure, partnership status, employment status, government regions, and technical variables (as a single component). In the sensitivity analysis, we additionally adjusted for smoking status, moderate physical activity, and body mass index. Variance inflation factors were <10. Total sample size was n=3307.

**Figure S7:** Association between environmental domains and epigenetic clocks in the main analysis and after multiple imputation. Weighted linear regression models were fitted separately for each exposure, expressed per interquartile range (IQR) increase. Models adjusted for age, sex, wave of data collection, income quartile, born in the UK, housing tenure, partnership status, employment status, government regions, and technical variables (as a single component). In the sensitivity analysis, missing data was imputed using multiple imputation by chained equations for 10 datasets and pooled based on Rubin’s rule. Sample size is n=3307 for the main, and n=3613 for the sensitivity analysis.

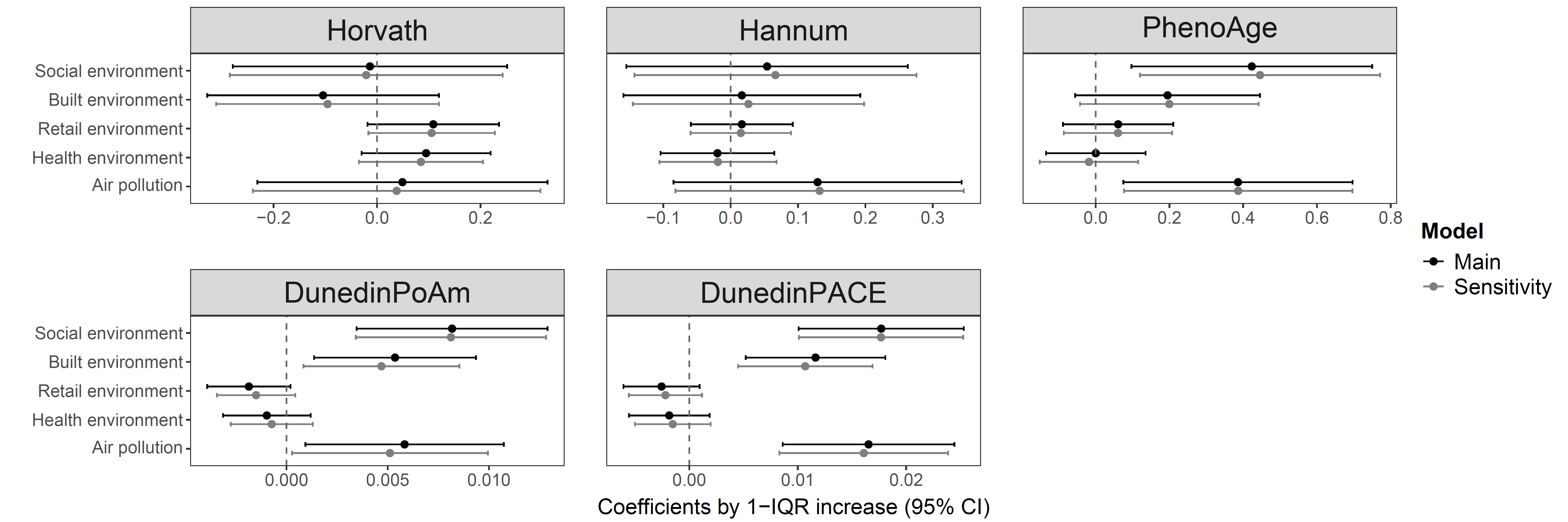

**Table S12**: Associations between environmental domains and epigenetic clocks, mutually adjusted domains.

| **Exposures** | **Main model** | | | |
| --- | --- | --- | --- | --- |
|  | b | 95% CI | p | p_FDR_ |
| Horvath clock |  |  |  |  |
| Social environment | 0.034 | -0.308, 0.375 | 0.846 | 0.846 |
| Built environment | -0.203 | -0.542, 0.135 | 0.239 | 0.399 |
| Retail environment | 0.121 | -0.080, 0.321 | 0.239 | 0.399 |
| Health environment | 0.029 | -0.175, 0.233 | 0.780 | 0.846 |
| Air pollution | 0.366 | -0.026, 0.759 | 0.067 | 0.337 |
| Hannum clock |  |  |  |  |
| Social environment | 0.024 | -0.245, 0.294 | 0.861 | 0.861 |
| Built environment | -0.153 | -0.427, 0.120 | 0.272 | 0.356 |
| Retail environment | 0.098 | -0.023, 0.218 | 0.113 | 0.295 |
| Health environment | -0.080 | -0.226, 0.067 | 0.285 | 0.356 |
| Air pollution | 0.256 | -0.065, 0.577 | 0.118 | 0.295 |
| PhenoAge |  |  |  |  |
| Social environment | 0.380 | -0.061, 0.820 | 0.091 | 0.151 |
| Built environment | -0.146 | -0.569, 0.277 | 0.499 | 0.624 |
| Retail environment | 0.181 | -0.024, 0.385 | 0.084 | 0.151 |
| Health environment | 0.002 | -0.209, 0.213 | 0.985 | 0.985 |
| Air pollution | 0.492 | 0.043, 0.942 | 0.032 | 0.151 |
| DunedinPoAm |  |  |  |  |
| Social environment | 0.007 | 0.001, 0.013 | 0.023 | 0.058 |
| Built environment | 0.002 | -0.004, 0.009 | 0.449 | 0.561 |
| Retail environment | -0.003 | -0.006, 0.000 | 0.069 | 0.115 |
| Health environment | 0.004 | 0.001, 0.007 | 0.023 | 0.058 |
| Air pollution | 0.001 | -0.006, 0.008 | 0.790 | 0.790 |
| DunedinPace |  |  |  |  |
| Social environment | **0.014** | **0.004, 0.024** | **0.006** | **0.031** |
| Built environment | 0.002 | -0.009, 0.013 | 0.754 | 0.754 |
| Retail environment | -0.002 | -0.008, 0.003 | 0.423 | 0.529 |
| Health environment | 0.006 | 0.000, 0.012 | 0.046 | 0.077 |
| Air pollution | 0.012 | 0.001, 0.024 | 0.040 | 0.077 |

Weighted linear regression models were fitted for each outcome, expressed per interquartile range (IQR) increase. The main models adjusted for age, sex, wave of data collection, income quartile, born in the UK, housing tenure, partnership status, employment status, government regions, and technical variables (as a single component). Variance inflation factors were <11. Total sample size was n=3307.

**Figure S8:** Determining the ideal number of environment clusters based on the a) elbow, b) silhouette, and c) gap methods. Sample size was n=3307.

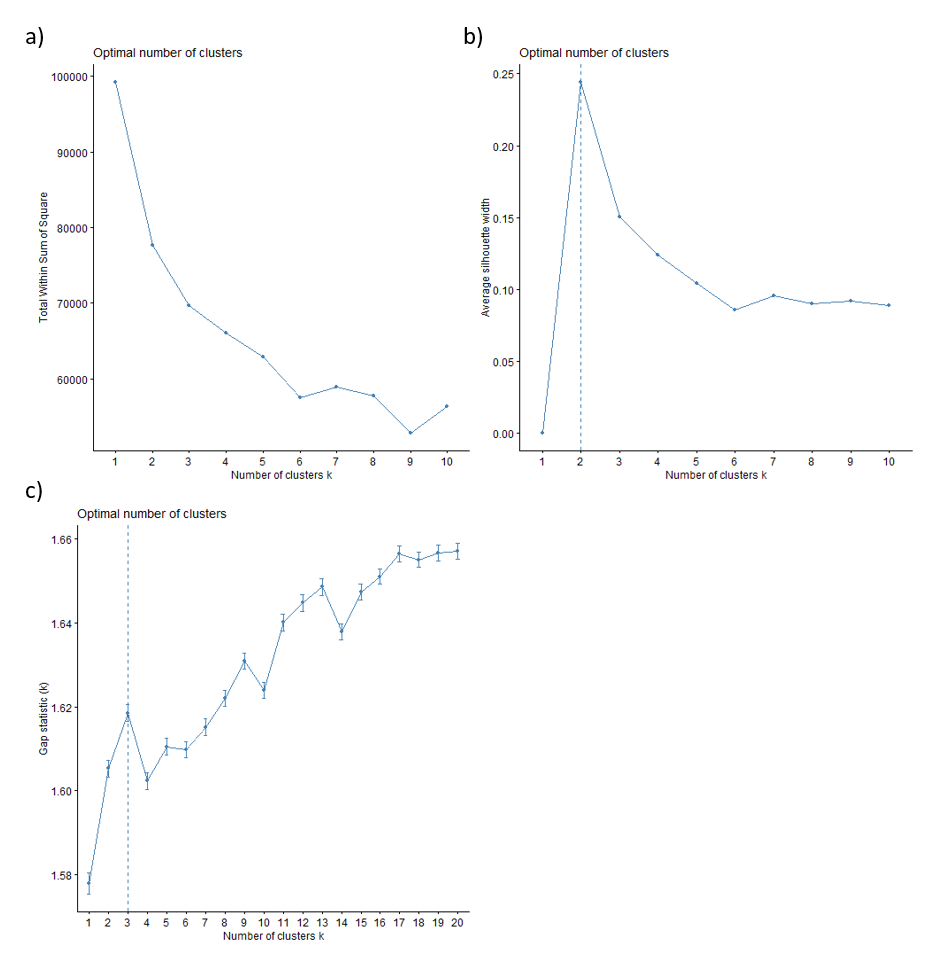

**Table S13**: Environmental clusters (two cluster solution) and epigenetic clocks, main models and sensitivity analyses.

| **Exposures** | **Main model** | | | **Sensitivity 1** | | | **Sensitivity 2** | | |
| --- | --- | --- | --- | --- | --- | --- | --- | --- | --- |
|  | b | 95% CI | b | b | b | p | b | b | p |
| Horvath |  |  |  |  |  |  |  |  |  |
| Urban (ref Rural) | -0.385 | -0.760, -0.011 | 0.044 | -0.376 | -0.748, -0.003 | 0.048 | -0.377 | -0.745, -0.009 | 0.045 |
| Hannum |  |  |  |  |  |  |  |  |  |
| Urban (ref Rural) | -0.100 | -0.386, 0.187 | 0.495 | -0.099 | -0.386, 0.188 | 0.498 | -0.068 | -0.351, 0.216 | 0.640 |
| PhenoAge |  |  |  |  |  |  |  |  |  |
| Urban (ref Rural) | -0.069 | -0.515, 0.378 | 0.764 | -0.124 | -0.555, 0.307 | 0.573 | -0.05 | -0.491, 0.391 | 0.823 |
| DunedinPoAm |  |  |  |  |  |  |  |  |  |
| Urban (ref Rural) | 0.008 | 0.001, 0.014 | 0.020 | 0.005 | 0.000, 0.009 | 0.042 | 0.007 | 0.001, 0.013 | 0.033 |
| DunedinPace |  |  |  |  |  |  |  |  |  |
| Urban (ref Rural) | 0.011 | 0.000, 0.022 | 0.043 | 0.008 | -0.001, 0.017 | 0.078 | 0.011 | 0.001, 0.022 | 0.038 |

Weighted linear regression models were fitted separately for each exposure-outcome combination, expressed per interquartile range (IQR) increase. The main models adjusted for age, sex, wave of data collection, income quartile, born in the UK, housing tenure, partnership status, employment status, government regions, and technical variables (as a single component). In sensitivity analysis 1, we additionally adjusted for smoking status, moderate physical activity, and body mass index. In sensitivity analysis 2, we imputed missing data using multiple imputation by chained equations for 10 datasets and pooled based on Rubin’s rule. Sample size is n=3307 for the main and sensitivity 1, and n=3613 for the sensitivity 2 analyses.

**Figure S9:** Weigthed mean exposure to environmental factors across urban, rural and mixed clusters. Values are presented on their original scales as weighted means with 95% confidence intervals. Area deprivation, crime and geographic accessibility are expressed as deciles, with higher values indicating greater deprivation, crime and accessibility. Social cohesion is a composite scale, with higher values indicating greater cohesion; population density is expressed as population per hectare; greenness is measured using the Normalised Difference Vegetation Index (NDVI); and distance to green and blue space is expressed in metres. Walkability is a composite index, light at night is measured in radiance, and grey space represents the percentage of built-up land cover. Distances to fast-food outlets, pubs, off-licences, tobacconists, gambling outlets, GPs, hospitals, dentists, pharmacies and leisure services are expressed in kilometres. Road traffic noise is categorised into decibel classes (1: ≤54.9 dB; 2: 55.0–59.9 dB; 3: 60.0–64.9 dB; 4: ≥65.0 dB). PM_2.5_, PM_10_, NO_2_, O_3_ are expressed in µg m^-3^, and temperature is expressed in degrees Celsius. Sample sizes were n=387 for rural, n=1423 for urban, and n=1497 for mixed.
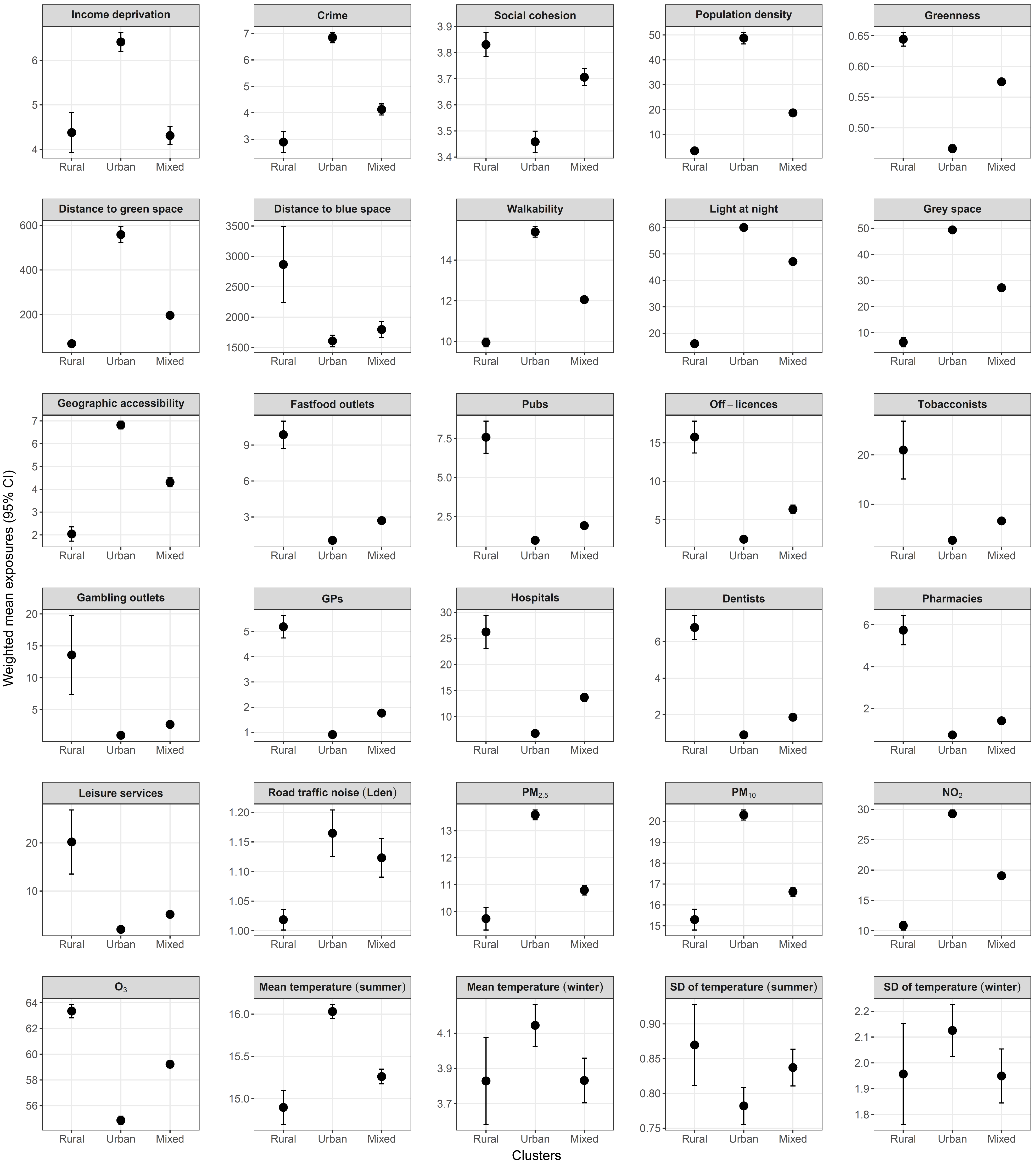

**Table S14**: Environmental clusters (three cluster solution) and epigenetic clocks, main models and sensitivity analyses.

| **Exposures** | **Main model** | | | **Sensitivity 1** | | |
| --- | --- | --- | --- | --- | --- | --- |
|  | b | 95% CI | p | b | 95% CI | p |
| Horvath |  |  |  |  |  |  |
| Mixed (ref Rural) | 0.081 | -0.465, 0.627 | 0.771 | 0.093 | -0.459, 0.645 | 0.742 |
| Urban (ref Rural) | -0.027 | -0.577, 0.524 | 0.924 | -0.021 | -0.577, 0.535 | 0.941 |
| Hannum |  |  |  |  |  |  |
| Mixed (ref Rural) | 0.151 | -0.260, 0.562 | 0.471 | 0.152 | -0.267, 0.571 | 0.476 |
| Urban (ref Rural) | 0.162 | -0.264, 0.588 | 0.455 | 0.149 | -0.284, 0.582 | 0.500 |
| PhenoAge |  |  |  |  |  |  |
| Mixed (ref Rural) | 0.366 | -0.251, 0.984 | 0.244 | 0.303 | -0.311, 0.917 | 0.333 |
| Urban (ref Rural) | 0.668 | 0.048, 1.288 | 0.035 | 0.498 | -0.115, 1.112 | 0.111 |
| DunedinPoAm |  |  |  |  |  |  |
| Mixed (ref Rural) | 0.004 | -0.006, 0.013 | 0.439 | 0.002 | -0.004, 0.009 | 0.496 |
| Urban (ref Rural) | 0.013 | 0.004, 0.023 | 0.008 | 0.008 | 0.000, 0.015 | 0.042 |
| DunedinPace |  |  |  |  |  |  |
| Mixed (ref Rural) | 0.005 | -0.012, 0.021 | 0.579 | 0.002 | -0.013, 0.016 | 0.824 |
| Urban (ref Rural) | 0.025 | 0.008, 0.042 | 0.005 | 0.016 | 0.001, 0.031 | 0.039 |

Weighted linear regression models were fitted separately for each exposure-outcome combination, expressed per interquartile range (IQR) increase. The main models adjusted for age, sex, wave of data collection, income quartile, born in the UK, housing tenure, partnership status, employment status, government regions, and technical variables (as a single component). In the sensitivity analysis, we additionally adjusted for smoking status, moderate physical activity, and body mass index. Total sample size was n=3307.

2.	Ministry of Housing Communities, and Local Government, Department for Levelling Up, Housing and Communities. English indices of deprivation. <https://www.gov.uk/government/collections/english-indices-of-deprivation>

3.	Welsh Government, StatsWales. Welsh Index of Multiple Deprivation. <https://statswales.gov.wales/Catalogue/Community-Safety-and-Social-Inclusion/Welsh-Index-of-Multiple-Deprivation>

4.	Scottish Government. Scottish Index of Multiple Deprivation 2020. <https://www.gov.scot/collections/scottish-index-of-multiple-deprivation-2020/>

5.	Shen Y, de Hoogh K, Schmitz O, et al. Europe-wide air pollution modeling from 2000 to 2019 using geographically weighted regression. *Environ Int*. Oct 2022;168:107485. doi:10.1016/j.envint.2022.107485

6.	DEFRA. Road Noise - Lden - England Round 2. Accessed 17.10.2025, <https://environment.data.gov.uk/dataset/2ec0abc3-ec1d-459c-aa6e-573672e94da7>

7.	Welsh Government. Environmental Noise Mapping 2012. Accessed 27.10.2025, <https://datamap.gov.wales/layergroups/inspire-wg:EnvironmentalNoiseMapping>

8.	Scottish Government. Scotland's noise. <https://noise.environment.gov.scot/index.html>

9.	Buckner JC. The development of an instrument to measure neighborhood cohesion. *American Journal of Community Psychology*. 1988;16(6):771-791. doi:<https://doi.org/10.1007/BF00930892>

10.	Daras K, Green MA, Davies A, Barr B, Singleton A. Open data on health-related neighbourhood features in Great Britain. *Scientific Data*. 2019/07/01 2019;6(1):107. doi:10.1038/s41597-019-0114-6
